# Data-driven neuropathological staging and subtyping of TDP-43 proteinopathies

**DOI:** 10.1101/2023.01.31.23285242

**Authors:** Alexandra L Young, Jacob W Vogel, John L Robinson, Corey T McMillan, Rik Ossenkoppele, David A. Wolk, David J. Irwin, Lauren Elman, Murray Grossman, Virginia M-Y Lee, Edward B Lee, Oskar Hansson

## Abstract

TAR DNA-binding protein-43 (TDP-43) accumulation is the primary pathology underlying several neurodegenerative diseases. Charting the progression and heterogeneity of TDP-43 accumulation is necessary to better characterise TDP-43 proteinopathies, but current TDP-43 staging systems are heuristic and assume each syndrome is homogeneous. Here, we use data-driven disease progression modelling to derive a fine-grained empirical staging system for the classification and differentiation of frontotemporal lobar degeneration due to TDP-43 (FTLD-TDP, n=126), amyotrophic lateral sclerosis (ALS, n=141) and limbic-predominant age-related TDP-43 encephalopathy neuropathologic change (LATE-NC) with and without Alzheimer’s disease (n=304). The data-driven staging of ALS and FTLD-TDP complement and extend previously described human-defined staging schema for ALS and behavioural variant frontotemporal dementia. In LATE-NC individuals, progression along data-driven stages was positively associated with age, but negatively associated with age in individuals with FTLD-TDP. Using only regional TDP-43 severity, our data driven model distinguished individuals diagnosed with ALS, FTLD-TDP or LATE-NC with a cross-validated accuracy of 85.9%, with misclassifications associated with mixed pathological diagnosis, age and genetic mutations. Adding age and SuStaIn stage to this model increased accuracy to 92.3%. Our model differentiates LATE-NC from FTLD-TDP, though some overlap was observed between late-stage LATE-NC and early-stage FTLD-TDP. We further tested for the presence of subtypes with distinct regional TDP-43 progression patterns within each diagnostic group, identifying two distinct cortical-predominant and brainstem-predominant subtypes within FTLD-TDP and a further two subcortical-predominant and corticolimbic-predominant subtypes within ALS. The FTLD-TDP subtypes exhibited differing proportions of TDP-43 type, while there was a trend for age differing between ALS subtypes. Interestingly, a negative relationship between age and SuStaIn stage was seen in the brainstem/subcortical-predominant subtype of each proteinopathy. No subtypes were observed for the LATE-NC group, despite aggregating AD+ and AD-individuals and a larger sample size for this group. Overall, we provide an empirical pathological TDP-43 staging system for ALS, FTLD-TDP and LATE-NC, which yielded accurate classification. We further demonstrate that there is substantial heterogeneity amongst ALS and FTLD-TDP progression patterns that warrants further investigation in larger cross-cohort studies.

## Introduction

The accumulation of tar DNA-binding protein 43 (TDP-43) underlies a variety of syndromes, being the primary cause of many cases of frontotemporal dementia (FTD) and most cases of amyotrophic lateral sclerosis (ALS). More recently TDP-43 accumulation has been identified as a frequent feature of Alzheimer’s disease and advanced aging, a phenomenon described as limbic-predominant age-related TDP-43 encephalopathy neuropathological change (LATE-NC) ^1^. Neuropathological staging systems have been derived by neuropathologists for each proteinopathy that describe the stereotypical spatiotemporal progression pattern of TDP-43 accumulation^1–5^. However, the manual derivation of these staging systems limits their spatiotemporal resolution and relies on the assumption that there is a single common spatiotemporal progression of TDP-43 for each syndrome. This limitation is of particular importance given that the mapping between TDP-43 pathology and clinical syndrome is imperfect^6,7^.

FTD is clinically and pathologically heterogeneous^8^, and recent work has recognized systematic variation in pathological features of TDP-43 underlying FTD^9^. ALS also exhibits substantial clinical heterogeneity^10^; whilst ALS is primarily characterised as a neuromuscular disorder, approximately 20% of ALS cases also have cognitive impairment^11^, and/or a typical FTD-syndrome. The concurrence of ALS and FTD suggest the possibility of these disorders constituting two extremes of an ALS-FTD spectrum disorder (ALS-FTD)^12^, supported by observations that *C9orf72* mutations can lead to either phenotype, or a mix of both^13,14^.

LATE-NC is a recently proposed terminology describing the commonly observed phenomenon of TDP-43 deposition mostly within medial temporal lobe structures in elderly adults^1^. Unlike FTD and ALS, the clinical syndrome that is associated with LATE-NC (termed ‘LATE’) currently does not have a corresponding set of criteria for a clinical diagnosis.

Although LATE is associated with memory impairment, discriminating whether cognitive impairment is due to LATE rather than other conditions is challenging without neuropathological evidence, particularly since it commonly co-occurs with Alzheimer’s disease (AD), which is also associated with memory impairment. LATE-NC is defined neuropathologically by a stereotypical distribution of TDP-43 in older adults^1,4,5^. However, the definition and description of this common neuropathological phenomenon is still being debated, and the terms LATE and LATE-NC have not met full consensus^15^.

Much is still unknown about the origin, progression and variability of TDP-43 aggregation as a whole, as well as within respective neurodegenerative diseases. For example, while many argue that there are clear clinical and pathological distinctions denoting distinct entities, there remains debate surrounding whether late-stage LATE-NC can be discriminated from early stage frontotemporal lobar degeneration due to TDP-43 (FTLD-TDP) given overlap of regions that are affected^15,16^. Similarly, the regional origin of FTLD-TDP is still unknown, with both the amygdala^3^ and frontoinsular or anterior cingulate^17^ proposed as candidate pathological epicentres.

A potential solution to capture the progression and spatial heterogeneity of TDP-43 proteinopathies is the use of data-driven disease progression modelling, which facilitates the probabilistic reconstruction of sequential disease progression patterns from cross-sectional data in a similar manner to traditional staging schema. However, in contrast to the traditional method of constructing neuropathological staging systems by hand, disease progression modelling can account for uncertainty in the level of pathology in each region and can enable the reconstruction of more complex progression patterns with large numbers of regions^18^. The Subtype and Stage Inference (SuStain) algorithm^19^ combines disease progression modelling with clustering to further enable the identification of subgroups of individuals (subtypes) with distinct disease progression patterns. This enables the probabilistic assignment of an individual to a subtype based on their subgroup, and stage based on their position along the inferred progression pattern for that subgroup. While previously applied to imaging datasets^19–22^, SuStaIn has recently been adapted to accommodate scored data often used in pathological assessment^23^.

Here we use SuStaIn to assess the progression and heterogeneity of TDP-43 accumulation in FTLD-TDP, ALS and LATE across 21 brain regions. We first estimate a single progression pattern for each group to enable comparison with existing TDP-43 staging schema. Next, we demonstrate the potential utility of disease progression models for the three-way classification of FTLD-TDP, ALS and LATE-NC. We further explore whether there is heterogeneity in the progression of TDP-43 within FTLD-TDP, ALS and LATE-NC by modelling multiple progression patterns within each group, and we test for differences between the resulting subgroups.

## Methods

### Dataset

Neuropathological samples with TDP-43 pathology were taken from the Center for Neurodegenerative Disease Research (CNDR) Brain Bank at the University of Pennsylvania, for which the tissue preparation, staining and immunohistochemistry procedures have been described previously^24^. Both sporadic cases and mutation carriers were included (see Table 1, S2). For each brain, up to 21 regions were sampled comprising the amygdala, hippocampal dentate gyrus, hippocampal cornu ammonis (CA)/subiculum, entorhinal cortex, anterior cingulate, superior/middle temporal gyrus, middle frontal gyrus, angular gyrus, occipital cortex, thalamus, globus pallidus, caudate/putamen, substantia nigra, midbrain, locus coeruleus, upper pons, cerebellum, medulla, orbitofrontal cortex, motor cortex and spinal cord (cervical spinal cord α-motoneurons in lamina 9) (**Figure S1**). Each region was assigned a score according to a semi-quantitative rating scale based on the extent of TDP-43 inclusions, where 0 = non-detectable, 0.5 = sparse, 1 = mild, 2 = moderate, and 3 = severe. The reliability of these scores was verified by independent expert neuropathologists. All individuals with any evidence of TDP-43 pathology in the brain were included in this study.

**Table 1.** Demographic information for all individuals included in this study, stratified by TDP-43 diagnosis. “LATE-AD-” = non-Alzheimer’s individuals with amygdalar but not medullar TDP-43 pathology. ‘‘Copathology” = Demonstrates at least one secondary copathology (other than LATE). AD = Alzheimer’s Disease; FTDS = Frontotemporal dementia spectrum; MND = Motor Neuron Disease; LBD = Lewy Body Disease; PMI = Postmortem Interval

|  | ALS | FTLD-TDP | LATE-AD+ | LATE-AD- |
| --- | --- | --- | --- | --- |
| N | 141 | 126 | 235 | 69 |
| Age at Onset | 59.5 (11.0) | 61.0 (9.0) | 70.2 (9.0) | 68.8 (10.0) |
| Age at Death | 63.7 (10.6) | 68.5 (10.0) | 80.9 (8.4) | 80.9 (8.7) |
| Disease Duration | 4.3 (4.3) | 7.3 (4.1) | 10.3 (4.3) | 11.9 (8.0) |
| % Female | 40.4% | 45.2% | 62.1% | 31.9% |
| Braak Stage | 1.7 (1.7) | 2.0 (1.6) | 5.8 (0.7) | 3.3 (1.4) |
| % APOE4 Carrier | 25.0% | 24.6% | 63.3% | 33.8% |
| % APOE2 Carrier | 12.9% | 18.2% | 5.2% | 8.8% |
| % Mutation Carrier | 19.9% | 46.8% | 4.2% | 5.8% |
| % Copathology | 76.6% | 88.9% | 42.7% | 55.1% |
| % AD Phenotype | 0.0% | 12.7% | 77.4% | 8.7% |
| % FTDS Phenotype | 1.4% | 81.7% | 9.4% | 15.9% |
| % MND Phenotype | 96.5% | 0.0% | 0.0% | 1.4% |
| % LBD Phenotype | 0.7% | 0.8% | 4.3% | 62.3% |
| % Other Phenotype | 1.4% | 4.8% | 9.0% | 11.6% |
| PMI | 12.6 (7.9) | 12.8 (7.3) | 12.6 (7.1) | 11.5 (6.7) |

Three data subsets – those with FTLD-TDP, ALS and LATE-NC – were extracted for disease progression modelling (**Table 1, S2**). The inclusion criteria for the FTLD-TDP and ALS subsets were a primary neuropathological diagnosis of FTLD-TDP^9,25^ and ALS^12^, respectively. Four ALS patients with a diagnosis of “ALS -Other”, which includes ALS without TDP-43 proteinopathy, and one ALS patient with a diagnosis of “ALS - Dementia” were excluded from this study. The inclusion criterion for the LATE-NC subset was a secondary or tertiary neuropathological diagnosis of LATE-NC^1^, with (LATE-AD+) or without (LATE-AD-) a primary neuropathological diagnosis of Alzheimer’s disease neuropathologic change^26^. A secondary neuropathological diagnosis of LATE-AD+ was made if individuals showed a primary diagnosis of AD (using NIA-AA criteria), did not have a secondary diagnosis of FTLD or ALS, and also had evidence of TDP-43^16^. The criteria for a neuropathological diagnosis of LATE-AD-were the absence of another TDP-43 neuropathological diagnosis (i.e. ALS, FTLD-TDP, or corticobasal degeneration), a TDP-43 score of 1 or more in the amygdala, and a TDP-43 score of less than 1 in the medulla (a region with near 100% sampling rate across diagnoses). These inclusion criteria were the sole criteria for inclusion in the study. In particular, we included individuals regardless of their TDP-type (A-E), and mutation carriers (e.g. *GRN, C9orf72*) and ALS-FTD cases were included. In total there were 126 individuals with FTLD-TDP, 141 with ALS and 304 with LATE-NC that were included in the SuStaIn modelling (**Table 1, S2**).

Age at death and estimated age at symptom onset were recorded for all individuals. Note that age of symptom onset refers to the age that primary symptoms were reported to manifest, which may or may not be symptoms directly related to TDP-43 neuropathology. Disease duration was calculated as the difference between age at symptom onset and age at death. Sex and interval between death and postmortem examination was also recorded for all patients. *APOE* genotype (ALS: 99%; FTLD-TDP: 100%; LATE-NC: 98%) and Braak tau stage (ALS: 95%; FTLD-TDP: 99%; LATE-NC: 97%) was also recorded for nearly all individuals using methods that have been previously described^24^. TDP-43 type was also available for FTLD-TDP patients using published criteria^9^. TDP-43 type was missing for two FTLD-TDP patients.

### Disease progression modelling using SuStaIn

Disease progression modelling was performed using Ordinal SuStaIn^23,27^. The SuStaIn algorithm combines clustering and disease progression modelling to estimate subtypes with distinct progression patterns from cross-sectional data. The Ordinal version of SuStaIn is specifically designed for modelling ratings data such as neuropathological ratings. Ordinal SuStaIn describes disease progression as a sequence in which different regions reach different scores, requiring as input the probability that a given score is observed in each region. In addition to learning the sequence in which different regions reach different scores, the algorithm quantifies the uncertainty in that sequence using Markov Chain Monte Carlo (MCMC) sampling. The uncertainty in the sequence is influenced by several factors including the number of samples available to estimate the sequence at different stages, the heterogeneity in the sequence between individuals, and the signal to noise ratio of the biomarker measurements (neuropathological ratings in this case). We modelled scores of 1, 2 and 3 in Ordinal SuStaIn, translating the scores of 0.5, 1, 2 and 3 to probabilities by evaluating a normal distribution around that score with a standard deviation of 0.5, and normalising by the sum of the probabilities of each score (**Table S1**). Note that, for example, a score of 0.5 (“sparse”) corresponds to an equal probability of the score being 0 or 1.

Missing neuropathological ratings were modelled as having an equal probability of having each score. Regions with a significant proportion of missing data (missing in more than 25% of subjects) were excluded when running Ordinal SuStaIn. For FTLD-TDP, the orbitofrontal cortex was excluded due to missingness, giving a total of 20 regions. For ALS, the occipital cortex, locus coeruleus and orbitofrontal cortex were excluded, with 18 regions remaining.

For LATE-NC, the motor cortex and spinal cord were excluded, leaving 19 regions. As each SuStaIn stage corresponds to a new region reaching a new score, there were N = 20*3 = 60 stages for FTLD-TDP, 18*3 = 54 stages for ALS, and N = 19*3 = 57 stages for LATE-NC. Ordinal SuStaIn was first used to estimate a single progression pattern for each group, enabling direct comparison with previous staging systems. Next, Ordinal SuStaIn was used to estimate multiple progression patterns for each group. The subtype progression patterns were cross-validated using 10-fold cross-validation and the optimal number of progression patterns (number of subtypes) within each group was chosen using the cross-validation information criterion, which balances model accuracy with model complexity^19^. For each progression pattern, individuals were first assigned to a subtype (in cases where n subtypes > 1), and were then assigned to a stage, based on maximum likelihood^19^. The SuStaIn stage can be thought of as a proxy for progression along a pathological trajectory. Subtypes were evaluated for cross-over events by plotting the distribution of subtype probability across stages. Subtype probability distributions that crossed chance levels (in this case, 50% for both ALS and FTLD-TDP given k=2 subtypes) after stage 1 were tagged as evidence for a crossover event. Crossover events suggest the possibility of discontinuity between variation observed before and after the event.

### Classification using SuStaIn

Three-way classification of FTLD-TDP, ALS and LATE-NC was initially performed by directly using the outputs of SuStaIn with a single progression pattern per group. SuStaIn was used to assign each individual to a group (FTLD-TDP, ALS or LATE-NC) and stage by evaluating the probability each individual belonged to each stage of each group’s progression pattern. The group and stage combination that had the highest probability was chosen as the predicted diagnosis for that individual. Individuals that had a most probable stage of 0 for their most probable SuStaIn-based diagnosis were labelled as “Unclassified”. Note that probabilities were computed out-of-sample for each individual (using 10-fold cross-validation when evaluating within-diagnosis), and took into account uncertainty in the progression pattern by integrating over the MCMC samples of the progression pattern output by SuStaIn. Unclassified stage 0 individuals were excluded before classification.

### Classification using logistic regression

A second predictive diagnostic model was evaluated using a combination of SuStaIn and logistic regression. Progression patterns for each group (ALS, FTLD-TDP, LATE-NC) were used to assess the probability that each individual fit within each progression pattern. Similar to above, these probabilities were calculated by evaluating the probability of the maximum likelihood stage within the progression pattern for each diagnosis. The three probabilities, along with maximum likelihood SuStaIn stage and age at death, were used as independent features in a multinomial logistic regression three-way classification model using an L2 penalty and Limited-memory BFGS (“lbfgs”) solver. The sample was divided into training (80%) and testing (20%) sets. In the training set, 10-fold cross-validation was used to optimise over a grid of 10 linearly spaced values between 1e-4 and 1e4 for the hyperparameter ‘C’. Performance was evaluated by recording accuracy, precision, recall and F1 score, both overall and class-weighted, of predictions in the left out test set. This process was repeated 100 times to establish confidence intervals around the measurements.

### Comparison of SuStaIn-derived progression patterns to previously described staging systems

Previously described staging systems were used to stage each individual based on regional presence of TDP-43 pathology. For ALS staging, we used the four-stage system described in ^2^. For FTLD-TDP, we used the four-stage system described in ^3^. Given that this system was derived based specifically on behavioural variant FTD (bvFTD) patients, we describe staging across both the whole group of FTLD-TDP patients, and across patients with a primary clinical diagnosis of bvFTD only. Since there are multiple proposed staging schemes for LATE-NC, we use three different proposed LATE-NC staging systems, as described in ^1^. For each staging system, composite regions of interest were created for each stage based on averaging the TDP-43 scores of the regions falling within that stage (see **Figure S2**). A patient was assigned to a stage if they showed a TDP-43 score equal to or greater than 1 for that stage as well as all previous stages in the regime. If a patient showed “out of order” staging (e.g. score >1 in stage I and III, but not II), they were assigned to an “Unclassifiable” category. Once all patients were staged using their group specific staging scheme, this stage was compared to the data-driven stage assigned to that patient. This approach allowed us to evaluate how much the data-driven SuStaIn stages resembled previously described manual staging systems of these various proteinopathies.

### Statistical analysis

Correlations were used to describe univariate relationships between SuStaIn stage and age, disease progression and total TDP-43 pathology, the latter calculated as the sum of all regional TDP-43 scores across an individual. Independent sample t-tests were used to assess differences between stage classifiable and unclassifiable patients (see “**Comparison of SuStaIn-derived progression patterns to previously described staging systems”**) in the probability of assignment to their a patient’s diagnostic progression pattern (see “**Classification using SuStaIn”**). Ordinary least squares (OLS) general linear models and χ^2^ tests were used compare misclassified to correctly classified patients from the machine learning classification analysis. To explore regional differences between subtypes, OLS general linear models were fit using each region as the dependent variable, subtype as the independent variable, and SuStaIn stage as a nuisance covariate (to ensure relationships were not due to differences in disease progression). Benjamini-Hochberg false discovery rate (FDR) was applied across all regions to correct for multiple comparisons. Multivariate logit maximum likelihood estimate regression models were used to investigate other subtype differences. Models were constructed using subtype as the dependent variable and the following as simultaneous independent variables: Age at symptom onset, age at death, sex, postmortem interval, *APOE* E2 carriage, *APOE* E4 carriage, Braak stage and SuStaIn stage. For the FTLD-TDP model only, TDP-43 type was also included as a covariate. Due to low numbers of individuals expressing TDP-43 type D (n=2), these individuals were excluded. In addition, two ALS individuals were assigned to “Stage 0” by SuStaIn, indicating they did not have sufficient TDP-43 pathology to be subtyped, and were excluded from the statistical analyses. Both the ALS (n=130) and the FTLD-TDP (n=115) models were fitted on a sample of subjects with complete data for all variables. To examine further clinical differences between ALS subtypes, another logit regression model was fit, this time using clinical symptom onset as the main independent variables, and all variables found to be significant in the previous analysis as covariates.

### Data availability

Source code for the Ordinal SuStaIn algorithm is available at https://github.com/ucl-pond/pySuStaIn. Data produced in the present study are available upon reasonable request to the authors.

## Results

### Progression of LATE-NC

Assuming a common progression pattern across all individuals with LATE-NC (**Figure 1**), SuStaIn identified that TDP-43 deposition was initially confined to the amygdala region (SuStaIn stages 1-2). This was expected given our definition of LATE-NC required TDP-43 pathology in the amygdala. During SuStaIn stages 3-9, TDP-43 deposition spread to the hippocampal CA/subiculum, followed by the entorhinal cortex and then to the hippocampal dentate gyrus. Subsequently, at SuStaIn stages 10-13, TDP-43 deposition emerged in the orbitofrontal cortex, followed by the anterior cingulate and then the superior/middle temporal gyrus. Moderate to severe TDP-43 pathology was present in all medial temporal regions before the appearance of TDP-43 pathology in cortical regions at SuStaIn stage 10. Beyond SuStaIn stage 13 there were few samples available, and the progression pattern had high uncertainty.

**Figure 1:**
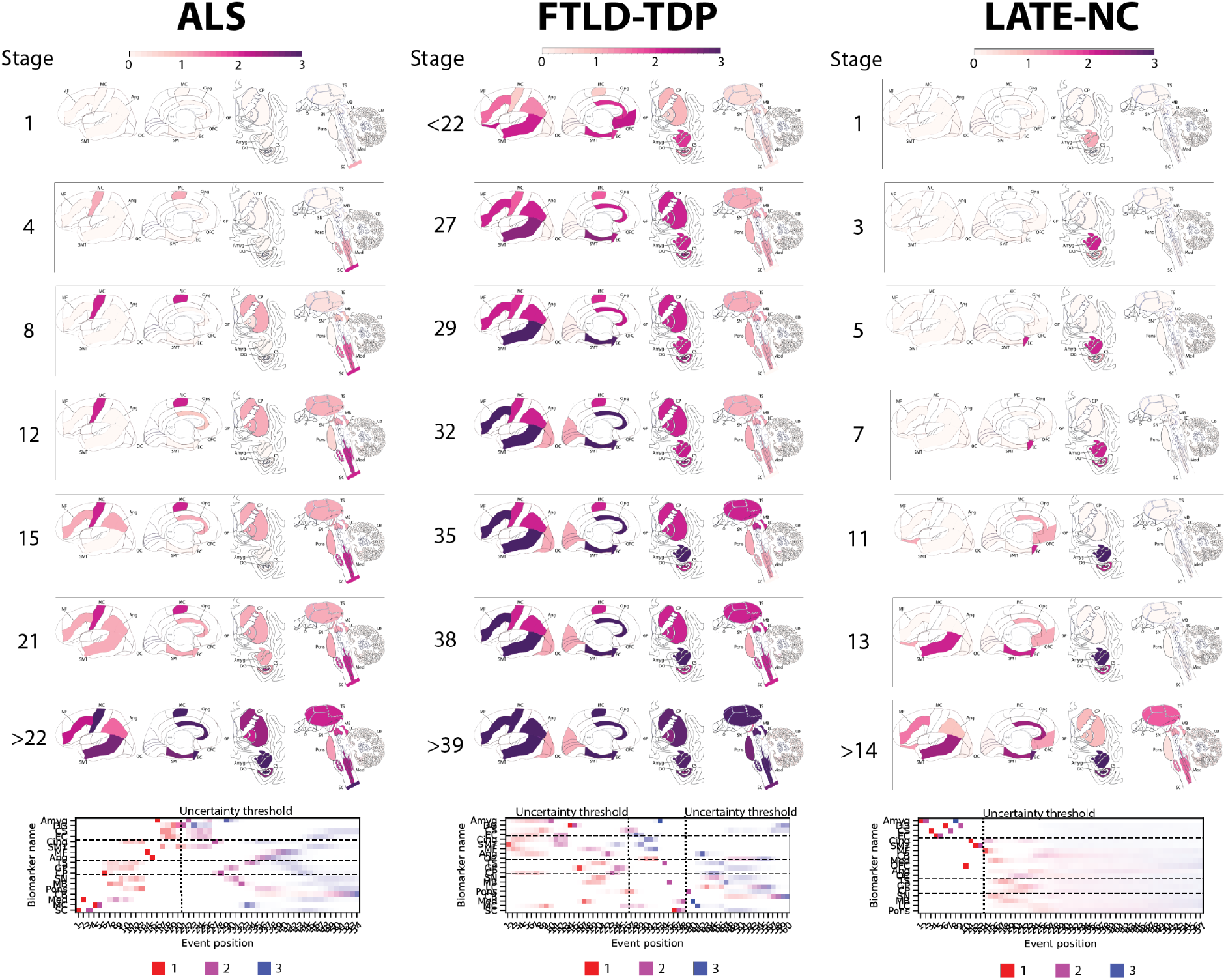
Progression patterns of TDP-43 proteinopathies. Inferred trajectory of regional TDP-43 progression based on individuals with a primary pathological diagnosis of ALS (left), a primary pathological diagnosis of FTLD-TDP (center), or a secondary or tertiary pathological diagnosis of LATE-NC. Colors within each brain region represent the cumulative sum of probabilities (0-1) summed across three stages of severity, light pathology (1) in red, moderate pathology (2) in purple, and severe pathology (3) in blue (see key at the bottom of each column). Only stages with reduced event ordering uncertainty (e.g. those represented by at least 5 individual donors) are shown. Below the brains are positional variance diagrams. Each box represents the degree of certainty that a given brain region (y-axis) has reached a given severity stage (red-light, purple-moderate, blue-severe) at a given SuStaIn stage (x-axis). Amyg = Amygdala; Ang = Angular Gyrus; CB = Cerebellum; Cing = Anterior Cingulate; CP = Caudate/Putamen; CS = CA1/Subiculum; DG = Dentate Gyrus; EC = Entorhinal Cortex; GP = Globus Pallidus; LC = Locus Coeruleus; MB = Midbrain; MC = Motor Cortex; MF = Middle Frontal Gyrus; Med = Medulla; OC = Occipital Cortex; OFC = Orbitofrontal Cortex; SC = Spinal Cord; SMT = Superior and Middle Temporal Gyrus; SN = Substantia Nigra; TS = Thalamus.

### Progression of ALS

Under the assumption of a single common progression pattern (**Figure 1**), SuStaIn estimated that TDP-43 deposition in ALS began in the spinal cord, before progressing to the medulla and motor cortex in SuStaIn stages 2-5. Subsequently, in SuStaIn stages 6-13, there was TDP-43 deposition in the caudate/putamen, thalamus, globus pallidus, midbrain, substantia nigra, pons and anterior cingulate. In SuStaIn stages 14-15, TDP-43 progressed to the middle frontal and angular gyri. By SuStaIn stage 21, TDP-43 was found in all regions sampled except the cerebellum, including regions of the medial temporal lobe.

### Progression of FTLD-TDP

Assuming a single common progression pattern for FTLD-TDP (**Figure 1**), SuStaIn identified that there was high uncertainty in the initial stages of TDP-43 progression in FTLD-TDP due to a lack of individuals with early stage TDP-43. The progression pattern was under-sampled and had high uncertainty before SuStaIn stage 27. While uncertainty was too high to draw confident inference, SuStaIn inferred that the amygdala, entorhinal cortex, anterior cingulate and superior/middle temporal gyrus were the first regions to display moderate TDP-43 pathology, between SuStaIn stages 10-13. When uncertainty became lower at SuStaIn stage 27, there was already moderate TDP-43 deposition throughout regions sampled from the medial temporal lobe, basal ganglia and cerebral cortex, with the exception of the occipital lobe, and light pathology throughout all subcortical regions sampled. At SuStaIn stages 28-32, TDP-43 deposition in the superior/middle temporal gyrus, anterior cingulate, entorhinal cortex, and middle frontal became severe, while mild TDP-43 deposition was seen in the occipital cortex. By SuStaIn stage 38, TDP-43 burden became severe in the amygdala, and moderate in the thalamus, midbrain, medulla, and also appeared in the spinal cord. Beyond SuStaIn stage 38, model uncertainty became too high for confident interpretation, though the locus coeruleus and cerebellum appeared to be the latest regions to be affected.

### Relationship between SuStaIn stage and age of onset, age at death and disease duration

Figure 2. shows the relationship between SuStaIn stage and total TDP-43 pathology, age of onset, age at death and disease duration in each condition. As expected and by design, SuStaIn stage was nearly collinear with total TDP-43 pathology, making it a convenient proxy for overall pathologic progression. FTLD-TDP had an inverse relationship between SuStaIn stage and global age of onset, age at death, and disease duration. In contrast the LATE-NC group had a positive relationship between SuStaIn stage and global age of onset and age at death, but no relation with disease duration. There was no significant relationship between SuStaIn stage and age or disease duration in ALS, however see ALS subtype results below.

**Figure 2:**
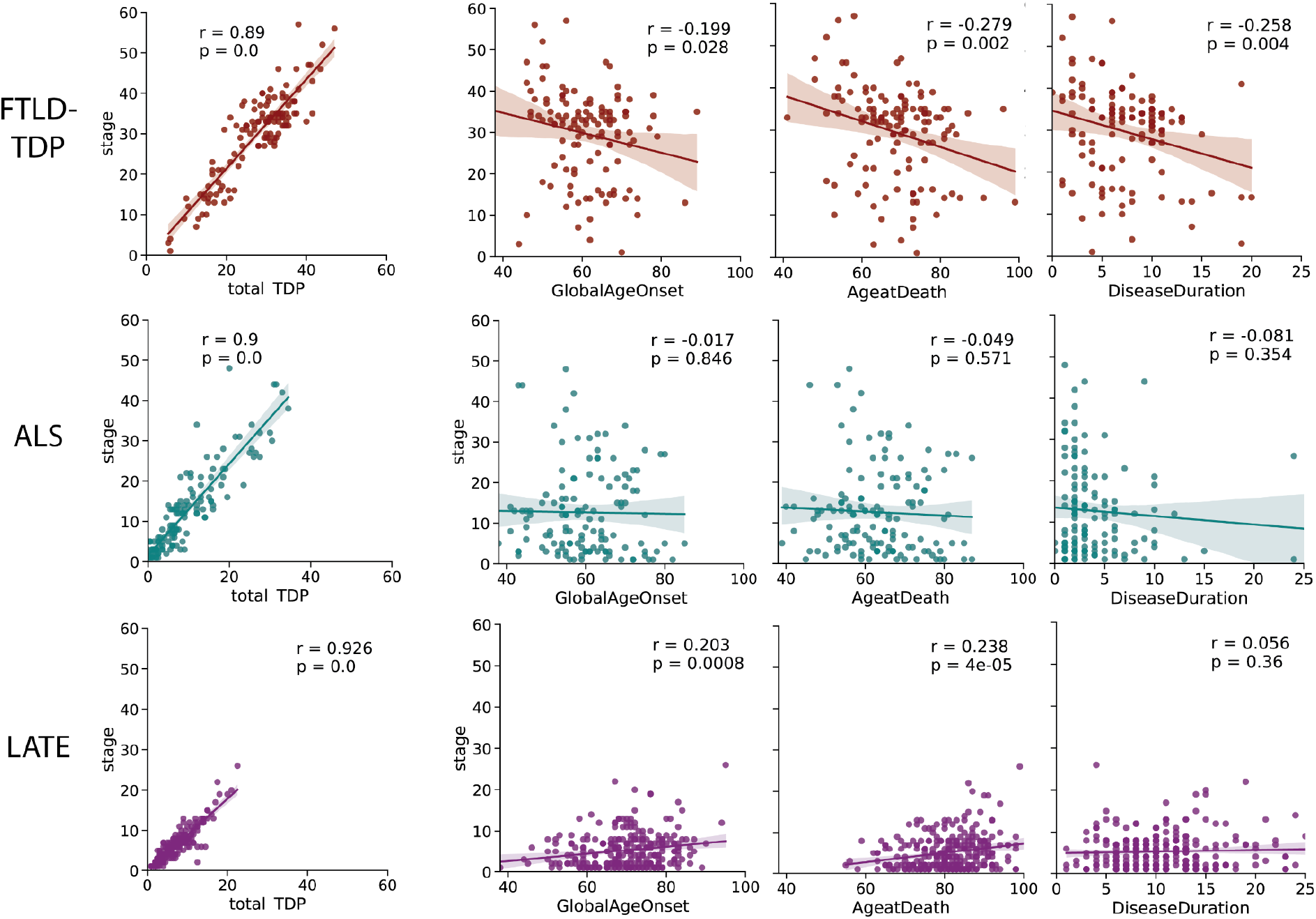
Relationships between age and disease progression. The relationship between total TDP-43 pathology, age of disease onset, age at death and disease duration with total TDP-43 pathologic burden (as measured with SuStaIn stage) across all three diagnostic categories. SuStaIn stage was strongly related with total TDP-43 pathology across all diagnostic groups. LATE-NC showed a significant positive relationship between SuStaIn stage and age (at onset and at death), whereas FTLD-TDP showed a significant negative relationship with age and disease duration.

### Comparison with previous staging schema

SuStaIn stage showed a good correspondence with existing staging schema (**Figure 3**), whilst providing additional granularity. Individuals were considered unclassifiable on existing staging schema if some regions had evidence of TDP-43 pathology (score greater than 1) that was consistent with a late stage, but other regions lacked evidence of TDP-43 pathology consistent with the earlier stages of the staging scheme. Notably, for all prior staging systems, unclassifiable individuals occurred at low, moderate and high levels of pathology.

**Figure 3:**
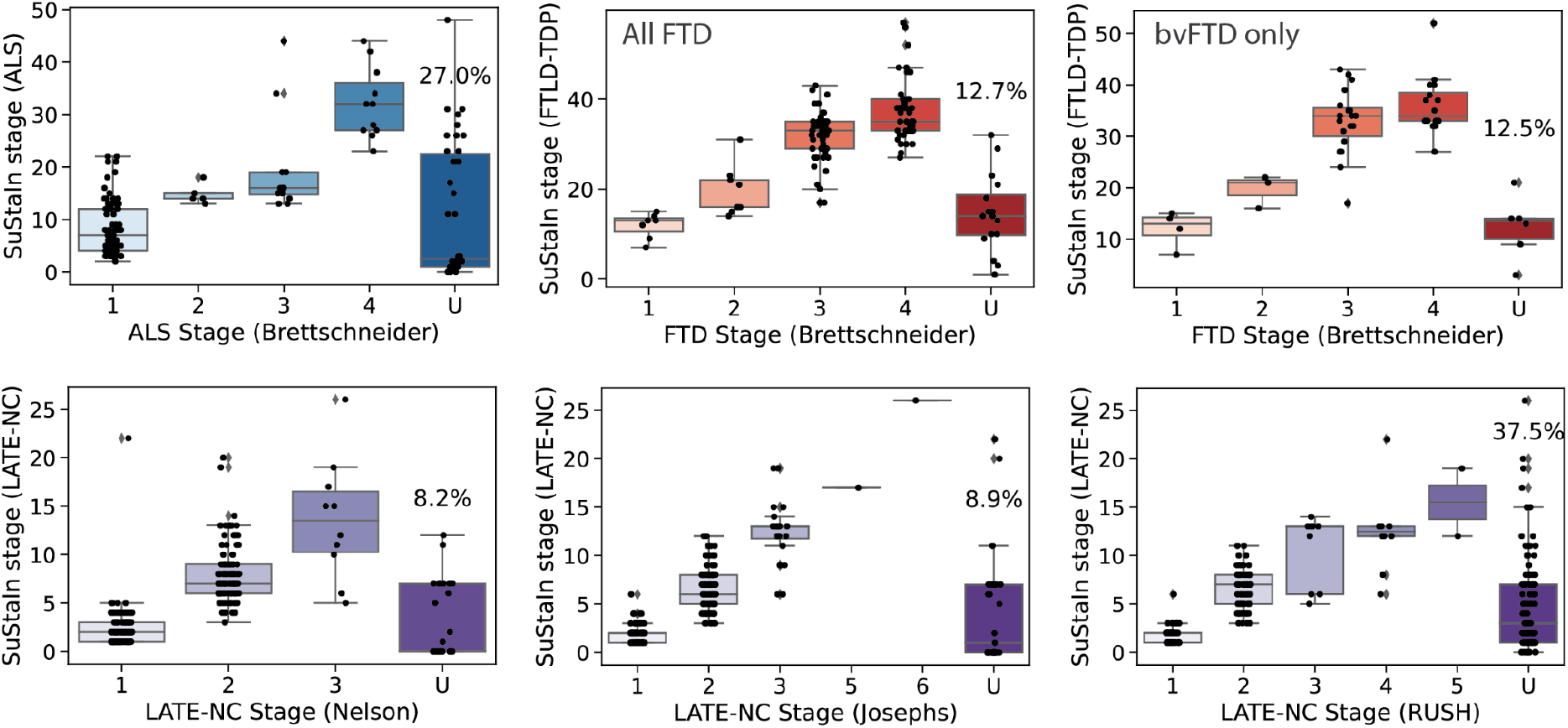
Comparison between SuStaIn-inferred TDP-43 pathological progression and previous staging systems. Individuals are staged based on various proposed staging criteria (refs; see **Figure S2**). TDP-43 pathology was averaged within regions belong to each stage. If an individual shows TDP-43 pathology (1+) in an advanced stage before showing TDP-43 pathology in a previous stage, they are considered to be “out-of-stage”, represented by “U” for “unclassifiable”. SuStaIn stage generally showed good correspondence with previous pathological staging systems.

Due to its probabilistic formulation, SuStaIn was able to stage individuals that were otherwise unclassifiable using existing staging schema. This probabilistic information could also be used to identify individuals that didn’t fit the staging system well. Indeed, compared to classifiable individuals, unclassifiable individuals had a lower (cross-validated) probability of belonging to the dominant progression pattern of their diagnostic group for the ALS staging system (t=2.60, p=0.01) and two of three LATE-NC staging systems (Nelson: t=1.46, p=0.14; Josephs: t=5.84, p<0.001; Rush: t=2.85, p=0.0046), though not for the FTLD-TDP staging system (full sample: t = -0.99, p=0.32; bvFTD only: t=-1.26, p=0.21).

### Three-way classification of FTLD-TDP, ALS and LATE-NC

Using SuStaIn, each participant was assigned a probability of belonging to the typical ALS, FTLD-TDP or LATE-NC progression (**Figure 4A-C**), regardless of TDP-type or genetic group. Most cases featured a high probability of their true pathological diagnosis. In fact, a winner-takes-all classification using only these probabilities resulted in a three-way classification accuracy of 85.9% (balanced accuracy = 81.3%; **Figure 4D, Table S3**), which was hampered most by a relatively low recall in FTLD-TDP diagnosis. Specifically, many FTLD-TDP cases were misclassified as LATE-NC (**Figure 4D**), and the probability of a LATE-NC diagnosis varied widely among FTLD-TDP cases (**Figure 4B**). After further optimising this model using logistic regression and including SuStaIn stage and age at death as features, out-of-sample three-way classification accuracy improved to 92.3% (balanced accuracy = 90.3%; **Figure 4E,F**; **Table S3**). Diagnostic tests of the decision boundaries suggested this improvement involved allowing higher ALS probability thresholds in the context of lower LATE-NC thresholds (**Figure S3A**). Most misclassifications across both the maximum likelihood and the logistic regression models involved FTLD-TDP patients misclassified as ALS or LATE-NC, and many of these misclassifications were high confidence (**Figure 4B**; **Figure S3A**).

Interestingly, cases having both ALS and FTLD-TDP as primary and secondary diagnosis (in either order) tended to have intermediate probabilities of both ALS and FTLD-TDP (**Figure S2B**). Many of the misclassifications were consistent with secondary underlying pathology, such as FTLD-TDP and ALS; for further case studies of misclassified individuals see Supplementary Note 1 and Figure S4.

**Figure 4:**
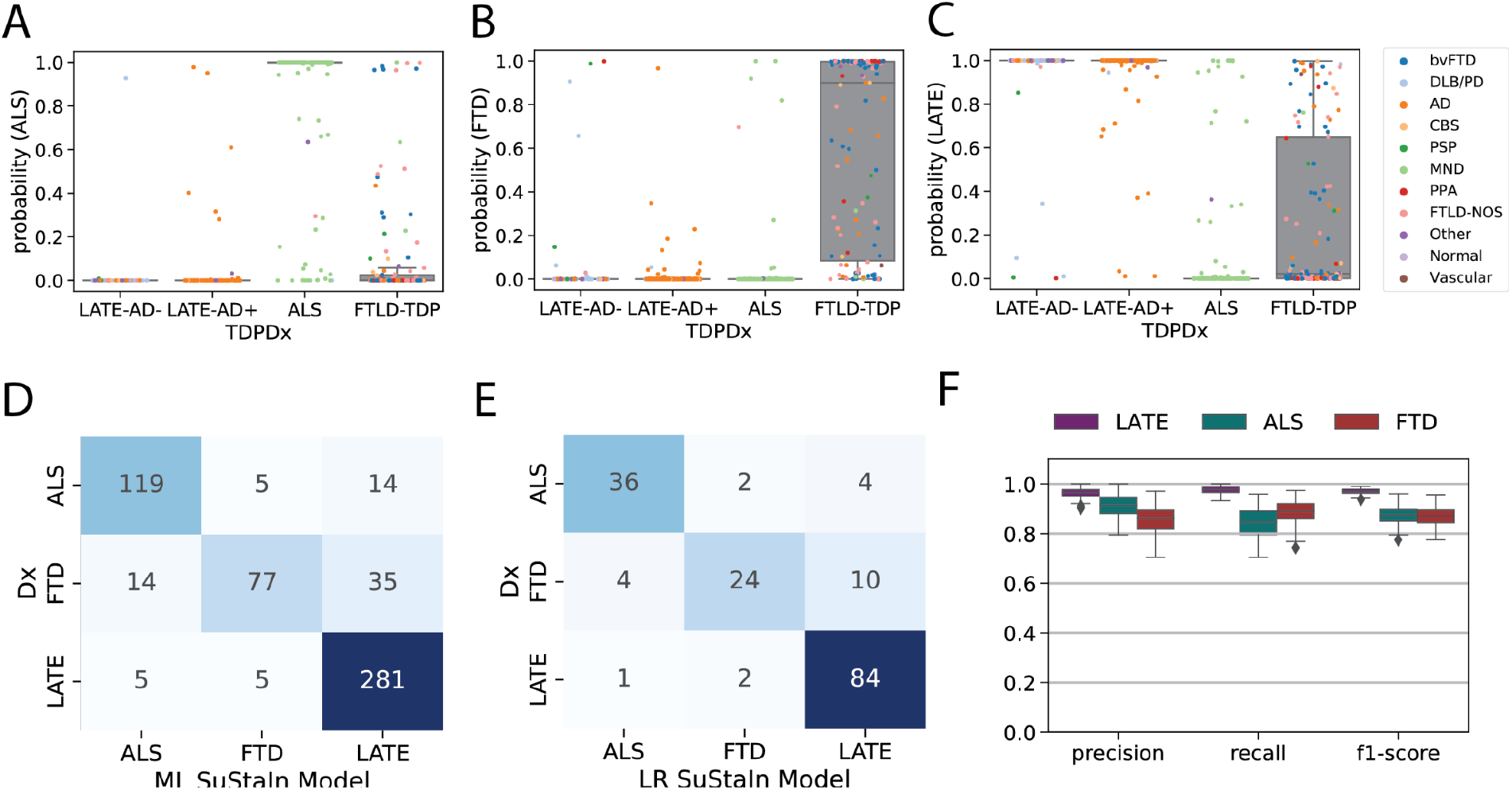
Pathology-based disease classification. All three diagnostic models were applied to all individuals regardless of TDP-type or genetic group, and the probability of the maximum likelihood stage was recorded. This probability represents a proxy for how well the individual’s regional TDP-43 pattern fit the model trajectory for LATE-NC/ALS/FTLD-TDP. Boxplots in (A-C) show the distribution of probabilities for each of the three diagnostic models (x-axis), stratified by pathological diagnosis (TDPDx on the x-axis), for (A) the ALS model, (B) the FTLD-TDP model and (C) the LATE-NC model. Note that each graph (A-C) includes a probability for every subject. Note also that probabilities were derived using 10-fold cross-validation for within-diagnosis assessments (e.g. ALS cases tested using the ALS model) to avoid over-fitting. Each individual is colored in accordance with their clinical diagnosis. Generally, individuals showed a high probability in models trained on their diagnosis, and a low probability in others. (D) A confusion matrix showing agreement between pathological diagnosis and maximum likelihood (ML) subtype model. True pathological diagnosis labels are represented on the y-axis, while predicted labels are shown on the x-axis. Individual pathological profiles tended to agree best with models fit to their clinical diagnostic group, but this was not true in all cases. (E) A logistic regression (LR) model was trained on the probabilities, plus ML SuStaIn stage and age at death, using 100 iterations of train/test splits. The confusion matrix shows the average agreement between pathological diagnosis and predicted subtype in the 100 left-out test groups. (F) Distribution of classification statistics for performance of the ML model across the 100 train/test splits, stratified by pathological diagnosis. See **Table S3** for further statistics and comparison with ML model.

### Differentiating LATE-NC from FTLD-TDP

Despite SuStaIn issuing good classification of LATE-NC from FTLD-TDP, the very low number of early stage FTLD-TDP cases or late-stage LATE-NC cases (**Figure 5A**) made it difficult to examine the point where these pathological entities could potentially intercept.

**Figure 5:**
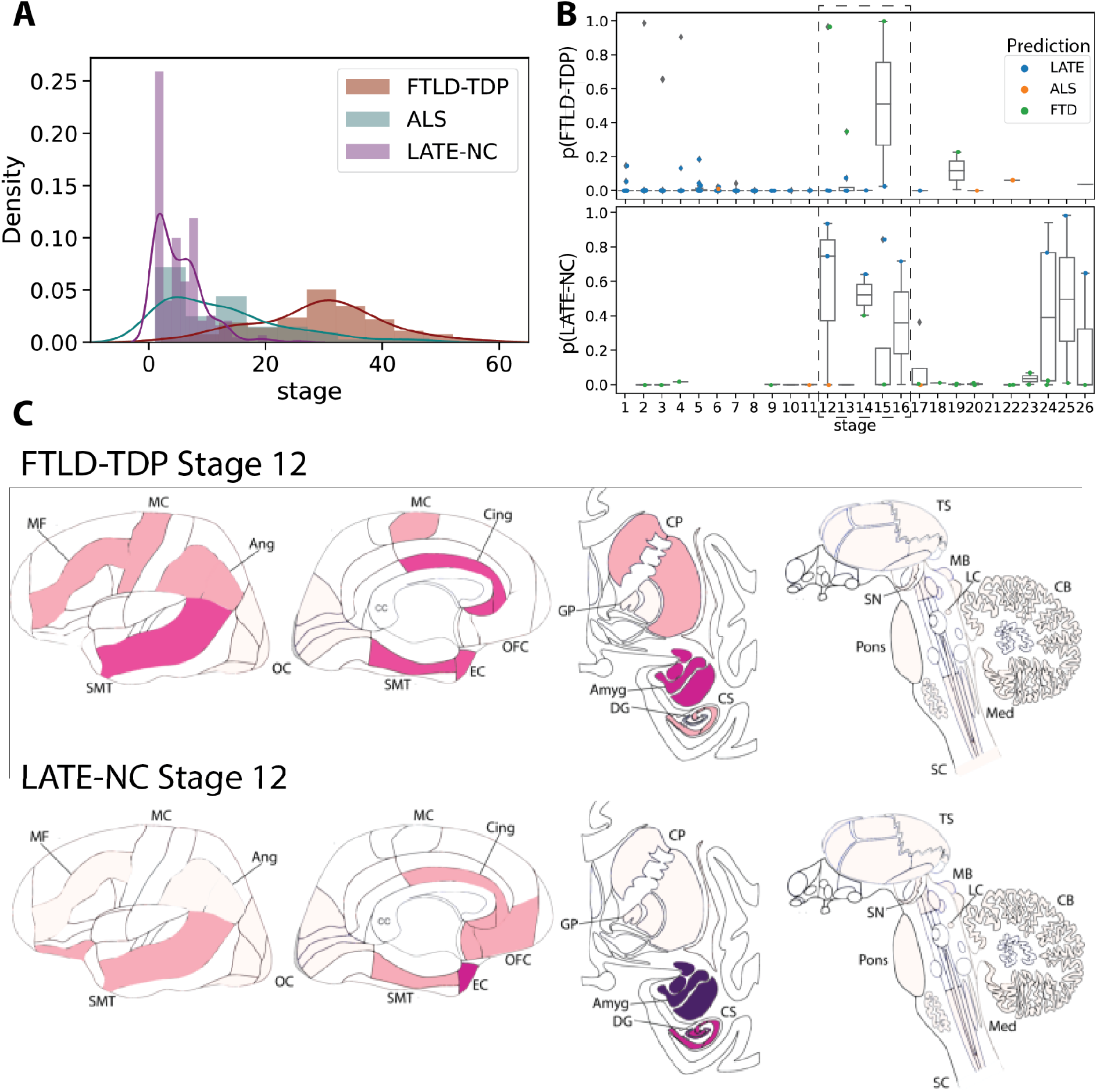
Comparing LATE-NC and FTLD-TDP. A) Histograms showing the proportion of individuals assigned to each stage, separately for FTLD-TDP, ALS and LATE-NC models/individuals. Nearly all LATE-NC individuals were assigned to stages 16 or lower, while nearly all FTLD-individuals were assigned to stages 16 or higher. B) SuStaIn tends to confuse LATE-NC and FTLD-TDP particularly the stages of the cross-over noted in A. The top plot represents the probability of being classified as FTLD-TDP among LATE-NC individuals, whereas the bottom plot represents the probability of being classified as LATE-NC for FTLD-TDP individuals. For both sets of patients, probability of misclassification is very low, except between stages 12-16. C) Comparison of FTLD-TDP and LATE-NC at Stage 12, with colors representing the amount of pathology using the same scale as Figure 1, ranging from white (no pathology) to red (light pathology) to purple (moderate pathology) to blue (severe pathology). See Figure 1 for abbreviations.

However, sufficient cases were present at SuStaIn stage 12 of both diagnostic groups to examine overlap of modelled SuStaIn’s progression patterns with a tolerable degree of certainty. Qualitatively, at stage 12, SuStaIn projects FTLD-TDP patients to have greater cortical burden of TDP-43 pathology, particularly in the anterior cingulate, as well as sparse TDP-43 burden in the basal ganglia. In contrast, stage 12 LATE-NC cases were projected to have greater (more severe) TDP-43 burden in the medial temporal lobe (**Figure 5C**).

However, among LATE-NC cases, a brief increase of FTLD probability was seen around LATE-NC stages 12-16, and a similar phenomenon of higher LATE-NC probability was seen in FTLD-TDP cases also around stages 12-16 (**Figure 5B**), indicating some degree of overlap between these two entities around these stages.

### Heterogenous pathological progression patterns of TDP-43 proteinopathies

For each neuropathologic diagnostic category, SuStaIn was refit allowing estimation of multiple progression patterns. For ALS, SuStaIn identified two subtypes of TDP-43 progression (**Figure 6**; **S5; S6**). The first subtype, which we labelled as subcortical predominant, had greater TDP-43 deposition in brainstem and subcortical regions, namely in the medulla, pons, basal ganglia, midbrain and substantia nigra. The second subtype, which we labelled as corticolimbic predominant, had greater TDP-43 deposition in cortical and medial temporal regions, namely the amygdala, hippocampus, superior/middle temporal gyrus, entorhinal cortex, and the occipital cortex (**Figure 6**; S6).

No evidence was found for putative crossover events between the two subtypes after stage 1 (**Figure S5**).

**Figure 6:**
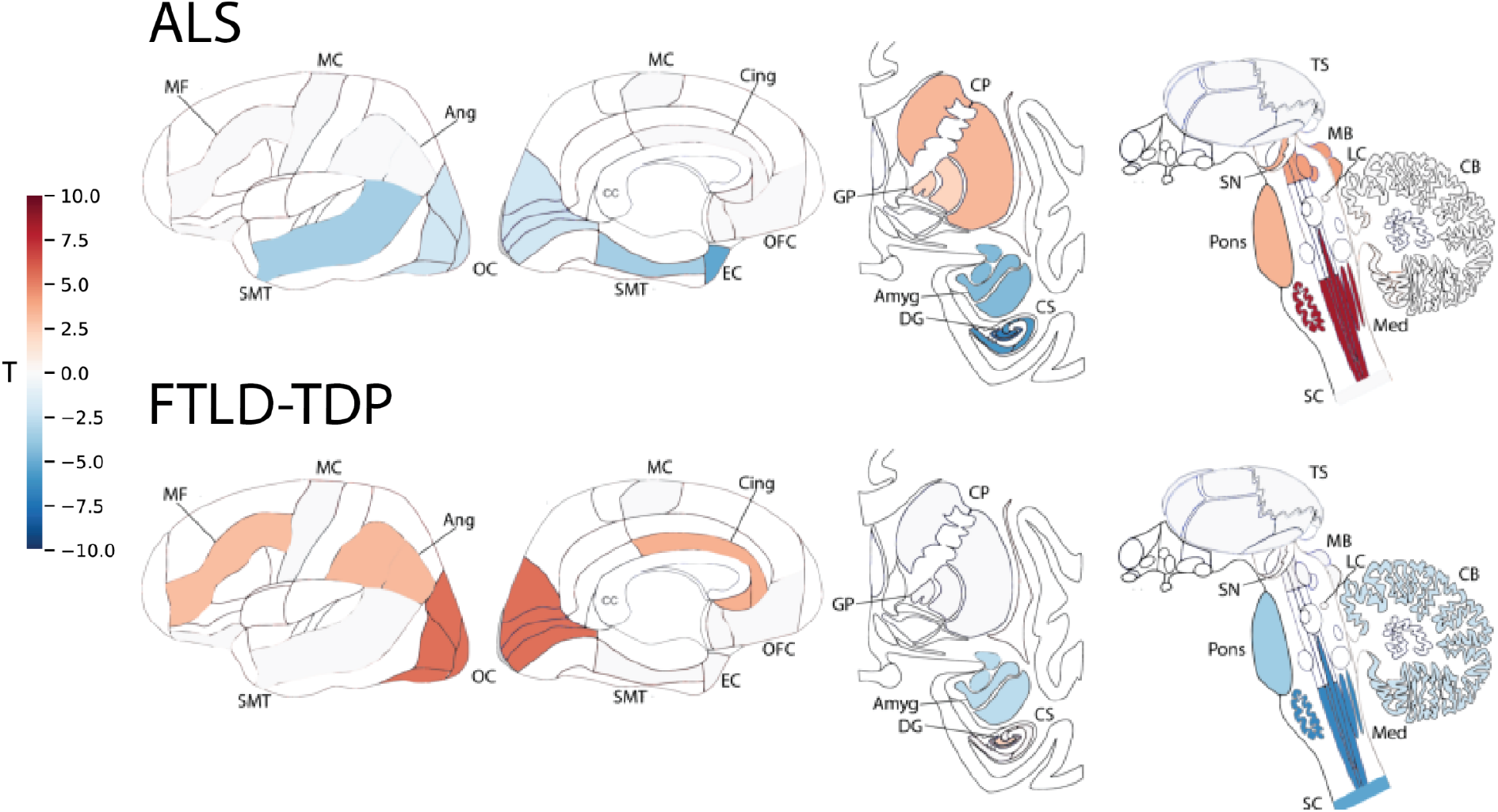
Subtypes of primary TDP-43 proteinopathies. T-maps showing regions that are significantly different between ALS subtypes (top) and FTLD-TDP subtypes (bottom), after controlling for SuStaIn stage and multiple comparisons. A positive t-value (red) indicates more severe TDP-43 pathology in Subtype 1 and a negative t-value (blue) indicates more severe TDP-43 pathology in Subtype 2. See Figure 1 for abbreviations.

When allowing SuStaIn to estimate multiple progression patterns for FTLD-TDP, SuStaIn identified two subtypes with distinct TDP-43 progression (**Figure 6**; **S5; S7**). The first subtype, which we labelled as cortical predominant, had greater TDP-43 deposition in the angular gyrus, occipital cortex, superior/middle temporal gyrus, middle frontal gyrus, anterior cingulate and dentate gyrus. The second subtype, which we labelled as brainstem predominant, had greater TDP-43 deposition in the medulla, pons, spinal cord, cerebellum and amygdala (**Figure 6; S7**). No evidence for a subtype crossover event was observed (**Figure S5**).

Despite aggregating a diverse set of individuals with and without Alzheimer’s disease, a single progression pattern best described the TDP-43 progression pattern for the n=304 participants with a secondary or tertiary neuropathological diagnosis of LATE-NC (**Figure S5)**.

### Clinical characteristics of FTLD-TDP subtypes

FTLD-TDP pathological subtypes were assessed for differences in demographic, pathological and genetic variables (**Table 2**). Compared to subtype 2 (brainstem predominant), subtype 1 (cortical predominant) participants were more likely to present with TDP-43 type C pathology, while subtype 2 was more likely to present with TDP-43 type B pathology and type E pathology, after adjusting for other covariates. There was also a significant interaction between SuStaIn subtype and SuStaIn stage on age at death (t=-2.85; p=0.005) and disease duration (t=-3.26, p=0.001), such that subtype 2 individuals showed a stronger negative relationship between SuStaIn stage and age at death (**Figure S8**).

**Table 2.** Comparison between regional TDP-43 subtypes of FTLD-TDP (top) and ALS (bottom). Each table represents the output of a single model including all covariates. S1 = subtype 1; S2 = subtype 2; PMI = post-mortem interval. ^a^ Stats not included because perfectly collinear when Age at Onset and Disease Duration both in model

| **FTLD-TDP** | **S1** | **S2** | **coef** | **p** | **vs B** | **p** | **vs C** | **p** | **vs E** | **p** |
| --- | --- | --- | --- | --- | --- | --- | --- | --- | --- | --- |
| **N** | 57 | 69 | - | - |  |  |  |  |  |  |
| **Age at Onset** | 60.5 (9.1) | 61.5 (9.0) | -0.00 | 0.89 |  |  |  |  |  |  |
| **Age at Death** | 68.3 (9.3) | 68.6 (10.7) | - | - |  |  |  |  |  |  |
| **Disease Duration** | 7.6 (2.8) | 7.1 (4.9) | 0.04 | 0.45 |  |  |  |  |  |  |
| **% Female** | 57.9% | 52.2% | 0.10 | 0.80 |  |  |  |  |  |  |
| **Braak Stage** | 2.0 (1.4) | 2.0 (1.8) | -0.13 | 0.58 |  |  |  |  |  |  |
| **% APOE4 Carrier** | 26.3% | 23.2% | 0.20 | 0.67 |  |  |  |  |  |  |
| **% APOE2 Carrier** | 17.5% | 18.8% | -0.21 | 0.70 |  |  |  |  |  |  |
| **% TDP-A** | 33.3% | 37.7% | - | - | 0.64 | 0.23 | -0.68 | 0.19 | 2.06 | 0.09 |
| **% TDP-B** | 24.6% | 34.8% | - | - |  |  | 1.32 | 0.03 | 1.42 | 0.24 |
| **% TDP-C** | 36.8% | 17.4% | - | - |  |  |  |  | 2.74 | 0.03 |
| **% TDP-E** | 1.8% | 8.7% | - | - |  |  |  |  |  |  |
| **SuStain Stage** | 30.1 (10.3) | 27.8 (11.3) | -0.03 | 0.13 |  |  |  |  |  |  |
| **PMI** | 12.4 (6.9) | 13.1 (7.6) | -0.01 | 0.84 |  |  |  |  |  |  |
| **ALS** | **S1** | **S2** | **coef** | **p** |  |  |  |  |  |  |
| **N** | 78 | 61 |  |  |  |  |  |  |  |  |
| **Age at Onset** | 57.4 (11.0) | 61.5 (10.2) | -0.04 | 0.09 |  |  |  |  |  |  |
| **Age at Death** | 62.0 (10.3) | 65.2 (10.3) | - | - |  |  |  |  |  |  |
| **Disease Duration** | 4.6 (4.5) | 3.9 (4.2) | 0.02 | 0.66 |  |  |  |  |  |  |
| **% Female** | 61.5% | 57.4% | 0.27 | 0.53 |  |  |  |  |  |  |
| **Braak Stage** | 0.8 (0.8) | 0.9 (0.9) | -0.06 | 0.82 |  |  |  |  |  |  |
| **% APOE4 Carrier** | 28.6% | 21.3% | 0.46 | 0.31 |  |  |  |  |  |  |
| **% APOE2 Carrier** | 15.6% | 9.8% | 0.66 | 0.38 |  |  |  |  |  |  |
| **SuStain Stage** | 10.9 (9.4) | 15.4 (10.7) | -0.05 | -0.01 |  |  |  |  |  |  |
| **PMI** | 14.3 (8.7) | 10.6 (6.1) | 0.07 | 0.02 |  |  |  |  |  |  |

Individuals with a secondary diagnosis of ALS were more likely to be subtype 2 (85.7% of 7 individuals with secondary neuropathological evidence of ALS vs 52.9% of 119 individuals without secondary neuropathological evidence of ALS), but this trend did not reach statistical significance (p=0.1928).

### Clinical characteristics of ALS subtypes

Similarly, differences in various clinical variables were tested between ALS subtype 1 (subcortical predominant) and ALS subtype 2 (corticolimbic predominant). Two confounding factors significantly discriminated between the two subtypes (**Table 2**): Subtype 1 had a longer postmortem interval, and also tended to die with less overall brain pathology (e.g. lower SuStaIn stage) compared to subtype 2. Covarying for these and other variables, there was a nonsignificant trend (p=0.09) for subtype 1 being associated with a younger onset age. There was a significant SuStaIn subtype x SuStaIn stage interaction on symptom onset age (t=2.56, p=.01) and age at death (t=2.72, p=0.007), where subtype 1 showed a stronger negative relationship (**Figure S8**).

## Discussion

In this study we employed data-driven disease progression modelling to derive an empirical system for staging and differentiating three major TDP-43 proteinopathies: ALS, FTLD-TDP and LATE-NC. The data driven staging schema derived from disease progression modelling corroborated previously described staging systems, whilst revealing additional detail in the progression patterns of each TDP-43 proteinopathy. Individuals diagnosed with ALS, FTLD-TDP or LATE-NC could be distinguished with high accuracy based on their probability of belonging to the learnt progression pattern for each proteinopathy. Most misclassified cases appeared to be driven by features of more than one TDP-43 diagnostic entity intermingling, as well as mutations and presence of co-pathologies. Our study further identified considerable heterogeneity in the progression of ALS and FTLD-TDP across individuals, but the progression of LATE-NC was found to be remarkably homogeneous.

Previous staging systems have been based on the delineation of a smaller, coarser set of stages based on the overall severity of pathology. In contrast, our data-driven staging systems were based on probabilistic inference of the sequential progression of TDP-43, enabling us to infer fine-grained progression patterns and to stage individuals probabilistically. Where previously described staging systems resulted in a substantial proportion of ‘unclassifiable’ individuals that didn’t precisely match the staging system, our data-driven method was able to probabilistically assign individuals to their best matching stage, accounting for the variability and uncertainty in the TDP-43 progression pattern for each particular TDP-43 proteinopathy. The probabilistic nature of the SuStaIn algorithm also allowed us to build proteinopathy classifiers by comparing an individual’s regional pattern to that of the dominant pattern of a given proteinopathy. This resulted in a robust and fully automated classification of diseases based only on an array of regional TDP-43 frequency scores plus age at death. The combined staging and classification could be useful for pathologists in classifying unusual cases and for establishing consistent quantification of individual TDP-43 progression for research studies.

Previous studies have described considerable pathological, clinical and biomarker heterogeneity in both ALS^28–30^ and FTLD-TDP^31,32^, and the two syndromes often co-occur within an individual^12^. Here we identified two major neuropathological subtypes in ALS and a further two in FTLD-TDP. The ALS subtypes broadly split into a subcortical predominant subtype with greater involvement of the spinal cord, medulla, pons and the caudate/putamen, and a corticolimbic predominant subtype with greater involvement of the temporal lobe, amygdala and hippocampus. Previous studies have similarly identified a subgroup of ALS cases with greater hippocampal and cortical involvement^33,34^. There is some controversy as to whether TDP-43 pathology in ALS originates in the motor cortex or the spinal cord. While our subtype analysis revealed subgroup differences in the relative timing of regional onset of motor cortex pathology, SuStaIn inferred the spinal cord as the first susceptible region for both ALS subtypes.

The FTLD-TDP patterns identified in this study divided into a brainstem predominant subtype with greater subcortical and brainstem involvement, and a cortical predominant subtype with greater cortical involvement. This is consistent with previous studies identifying subcortical involvement in FTD subgroups^35,36^, with the brainstem predominant subtype having greater subcortical involvement. The brainstem predominant subtype was more likely to present with TDP-43 type B and E pathology, whilst the cortical predominant subtype was more likely to present with TDP-43 type C pathology. This aligns with previous work demonstrating that TDP-43 type B is associated with FTD-ALS phenotypes^32,37–39^, with the subcortical and brainstem involvement in the brainstem predominant subtype being closer to the pattern of TDP-43 progression in ALS. As with ALS, further studies in larger samples may reveal further subdivision of the subtypes.

SuStaIn stage was positively correlated with age in LATE-NC, in agreement with previous studies^1^. However, in FTLD-TDP SuStaIn stage was found to be negatively correlated with age. Interestingly, when allowing for subtypes, both ALS and FTLD-TDP exhibited subtypes with an increased amount of subcortical pathology and an inverse relationship between age and pathology, suggesting that age associations may have been driven by subgroup associations for FTLD-TDP and negated by subgroup associations for ALS. The factors leading to positive and negative relationships between pathology and age are complex and incompletely understood. A positive association with age might be produced by a slow progressing (typically late onset) pathology, which is often present alongside other pathologies and is unlikely to be the primary cause of death. A negative association with age, in contrast, may be produced by a fast progressing (often early onset) pathology, with a higher burden of pathology being found in individuals with faster progression who consequently die at a younger age.This idea is supported by previous studies that have found inverse correlations between the primary pathology and age, such as between age and tau deposition in Alzheimer’s disease^20,40,41^. An alternative explanation for an inverse correlation between disease stage and age is that more neuronal death in older (or more progressed) subjects leads to lower proportions of TDP-43 pathology being stained on histopathological slides. Some evidence for this phenomenon is presented by individuals with long-lasting ALS, who demonstrate decreased TDP-43 staining due to neuronal death of TDP-43-vulnerable neurons^42,43^. One final explanation for an inverse correlation with age is that younger subjects may have higher neurocognitive reserve, and are therefore able to function under a greater burden of pathology^44^.

We identified a single progression pattern for LATE-NC, suggesting that the pattern of TDP-43 progression in LATE-NC is relatively stereotypical, or at least spread around a single major mode. We didn’t find evidence in the present study that the progression of LATE-NC differed between individuals with or without Alzheimer’s disease, although our sample was dominated by individuals with Alzheimer’s disease and larger studies will be required to confirm this. A recent study by Cykowski et al.^45^ identified subtypes of LATE-NC with distinct TDP-43 inclusion morphologies. Our study does not necessarily contradict this study as we didn’t consider the morphology of the TDP-43 inclusions when deriving subtypes and stages. It is also worth noting that we didn’t consider very early ‘sparse’ pathology, whereas early pathology was found to be more heterogeneous in the study by Cykowski et al.

The regions affected by LATE-NC and FTLD-TDP are known to overlap and the similarity of their TDP progression patterns has not yet been fully determined, although some studies suggest that they can be differentiated^16^. Whilst could differentiate LATE-NC and FTLD-TDP with high accuracy, it is important to note that there was a systematic under-sampling of individuals with early stages of FTLD-TDP and late stages of LATE-NC in our dataset. We did observe subtle differences for the few individuals at overlapping middle stages of FTLD-TDP or LATE-NC.. However, we also observed a clear decrease in the confidence of classifying these individuals, suggesting that these subtle differences may not be strong enough to differentiate one condition from the other at an individual level. Similarly, there was considerable uncertainty in the early stages of the FTLD-TDP progression pattern and the late stages of the LATE-NC progression pattern. There is some controversy as to whether FTLD-TDP and LATE-NC share similar origins, where FTLD-TDP may be an earlier-onset and far more aggressive variant of LATE-NC. Based on our observations, we can neither confirm nor reject this hypothesis, especially since early FTLD-TDP is undersampled in our cohort. Closer study of TDP-43 morphology in LATE-NC and FTLD-TDP may help to resolve this controversy.

Our study has a number of limitations that warrant further consideration in future work. The neuropathological sample was obtained from a tertiary academic centre and may have a referral bias. FTLD-TDP is also known to be highly heterogeneous, potentially with distinct progression patterns in carriers of genetic mutations for FTLD-TDP, however our study was underpowered to resolve mutation-specific patterns. The SuStaIn algorithm infers subtypes with distinct progression patterns from cross-sectional data, relying on the assumption that TDP-43 proteinopathies progress sequentially from region to region, as is the case in all neuropathological studies. The SuStaIn algorithm also assumes that each individual has a single TDP-43 proteinopathy (FTLD-TDP, ALS or LATE-NC), an assumption violated by several individuals in this study, leading to misclassification. Our study determined progression patterns using neuropathological samples from a large number of regions.

Whilst ideally suited to determining the progression pattern of each TDP-43 proteinopathy, this means that the derived staging systems may require sampling of a larger range of regions than is necessary to differentiate one proteinopathy from another. There are also several regions that were not sampled in the present study that merit further investigation in future studies. Our study only considered the total amount of TDP-43 pathology in each region and did not consider the specific TDP-43 inclusion morphologies, which may provide further stratification if considered in future work, e.g. TDP type may enhance the differentiation of FTLD-TDP and LATE-NC^46^ but was only available for a subset of individuals in our study. In this study, we focused on comparing SuStaIn to staging schema of predefined neuropathological groups, running SuStaIn separately in each group. While consistent with current neuropathological practice, this may bias our results towards only reproducing these preconceived diagnostic categories. For example, diagnosis of LATE-NC in this study required amygdalar TDP-43 pathology, which may bias the amygdala as the earliest region to show TDP-43 burden. A similar issue may be present with the spinal cord and ALS, though other criteria are used in the neuropathological definition of ALS as well.

Overall, we develop an empirical pathological staging system for ALS, FTLD-TDP and LATE-NC, using data-driven disease progression modelling. We demonstrate that this staging system is sufficient for staging and accurate classification and can provide probabilistic information that aids classification of individuals that deviate from the stereotypical progression pattern for each TDP-43 proteinopathy. Our results demonstrate that there is substantial heterogeneity amongst ALS and FTLD-TDP progression patterns, whilst we identify that LATE-NC is relatively homogeneous in this cohort. Data-driven disease progression modelling is a useful tool for neuropathological staging and management and can aid in unravelling neuropathological heterogeneity.

## Data Availability

Data produced in the present study are available upon reasonable request to the authors.

## Acknowledgements

We would like to acknowledge the late John Q. Trojanowski, MD. Ph.D., who also participated in study design and interpretation of results. ALY is supported by an MRC Skills Development Fellowship (MR/T027800/1). JWV acknowledges funding from the NIH (T32MH019112) and the SciLifeLab & Wallenberg Data Driven Life Science Program (grant: KAW 2020.0239). This work was supported by the Penn Udall Center of Excellence in Parkinson’s Disease Research, the Penn Center for Neurodegenerative Disease Research, and the Department of Pathology and Laboratory Medicine of the Penn Alzheimer’s Disease Center. The current work and other research at these centres were supported by the NIH (P30 AG072979, P01AG066597, U19AG062418). OH was supported by the Swedish Research Council (2016-00906), the Knut and Alice Wallenberg foundation (2017-0383), the Marianne and Marcus Wallenberg foundation (2015.0125), the Strategic Research Area MultiPark (Multidisciplinary Research in Parkinson’s disease) at Lund University, the Swedish Alzheimer Foundation (AF-939932), the Swedish Brain Foundation (FO2021-0293), The Parkinson foundation of Sweden (1280/20), the Cure Alzheimer’s fund, the Konung Gustaf V:s och Drottning Victorias Frimurarestiftelse, the Skåne University Hospital Foundation (2020-O000028), Regionalt Forskningsstöd (2020-0314) and the Swedish federal government under the ALF agreement (2018-Projekt0279).

