## Supplementary Material for "Data-driven neuropathological staging and subtyping of TDP-43 proteinopathies"

### Supplementary Note 1: Case studies of misclassified individuals

Misclassified individuals were identified by comparing each individual's neuropathological group (ALS, FTLD-TDP or LATE-NC) with their most probable SuStaIn-based diagnosis. Individuals that were labelled as SuStaIn stage 0 were excluded from the list of misclassified individuals. A case study of each misclassified individual was performed by a neuropathologist (EBL) to check for notable secondary or tertiary diagnoses or genetic mutations. These observations were tested statistically using ANOVAs or $\chi^2$ tests as appropriate to compare cases by their predicted diagnosis. Heatmaps were generated to qualitatively compare the regional pattern of misclassified cases to that of correctly classified cases. In addition, cases were identified as ALS-FTLD when both ALS and FTLD-TDP were listed as the primary and secondary neuropathological diagnosis (in either order). The SuStaIn-derived probability of classifying these subjects as ALS or FTLD-TDP was visualised compared to cases with a neuropathological diagnosis of ALS only, FTLD-TDP only, or neither.

In total there were 78 individuals that were misclassified by SuStaIn, and a further 15 were designated as "Unclassifiable" due to a lack of significant TDP-43 deposition (i.e. Stage 0). However, many of the misclassifications were consistent with secondary underlying pathology. Of 16 patients with both FTLD-TDP and ALS included in their neuropathological diagnosis, 12 (75%) were misclassified. Among 14 FTLD-TDP cases misclassified as ALS, five had a secondary neuropathological diagnosis of ALS, compared to 2 of 32 misclassified as LATE-NC and 0 of 80 correctly classified individuals ($\chi^2$ = 29.0, p = 5e-7). Furthermore, nine of the 14 had primary or secondary clinical diagnosis of ALS, compared to 4 of 32 misclassified as LATE-NC and 1 of 80 correctly classified ($\chi^2$ = 48.0, p = 3.7e-11, Tukey[ALS] p < 0.0001, Tukey[LATE-NC] p = 0.0028). Similarly, two of five ALS patients misclassified as FTLD-TDP had a secondary neuropathological diagnosis of FTLD-TDP, compared to 3 of 14 misclassified as LATE-NC and 4 of 119 correctly classified ALS patients ($\chi^2$ = 16.2, p = 0.0003, Tukey[FTLD] p = 0.0028, Tukey[LATE-NC] p = 0.013). In addition, in ALS patients misclassified as LATE-NC, 13 of 14 also had primary age-related tauopathy (PART) or AD neuropathologic change, compared to 2 of 5 misclassified as FTLD and 83 of 119 correctly classified ($\chi^2$ = 5.7, p = 0.059). However, no such trend was seen among FTLD-TDP patients misclassified as LATE-NC.

An examination of the regional patterns of misclassified cases supported the notion that these cases showed regional pathology patterns that shared features with multiple neuropathological diagnoses (**Figure S4**). For example, all four LATE-NC cases misclassified as ALS had TDP-43 pathology in the midbrain, and three also had medullar pathology. Similarly, all but one LATE-NC case misclassified as FTLD-TDP featured TDP-43 pathology in the anterior cingulate and the temporal lobe. These same trends were seen in ALS – most ALS cases misclassified as LATE-NC has substantial medial temporal lobe pathology, and cases misclassified as FTLD-TDP had frontal and both medial and lateral temporal lobe pathology.

Other measurable features appeared to contribute somewhat to misclassification of cases. For example, 50% (7 of 14) FTLD-TDP cases misclassified as ALS carried a *C9orf72* expansion, vs 25% (8 of 32) cases misclassified as LATE-NC and 21.3% (17 of 80) of correctly classified cases ($\chi^2$ = 5.2, p=0.074). Interestingly, three of seven LATE-NC cases misclassified as FTLD-TDP had a primary neuropathological diagnosis of progressive supranuclear palsy (PSP), compared to 0 of 4 misclassified as ALS and 11 of 280 correctly classified. All three cases exhibited diffuse cortical TDP-43 pathology, as well as light pathology in the basal ganglia (**Figure S4**). There was also a significant effect of SuStaIn stage across ALS (means by predicted diagnosis [sd]: ALS=10.9 [9.2], FTLD-TDP=22.8 [17.0], LATE-NC=23.2 [11.4], F = 12.8, p = 8e-06), FTLD-TDP (ALS=35.5 [13.9], FTLD-TDP=27.0 [10.3], LATE-NC=30.6 [9.8], F = 4.4, p = 0.014) and LATE-NC (ALS=18.5 [8.7], FTLD-TDP=9.7 [6.7], LATE-NC=4.8 [3.6], F = 31.6, p = 4e-13) cases, where misclassified individuals had a more advanced SuStaIn stage indicating more total pathology. Age at death also contributed to misclassification among LATE-NC (F = 5.9, p = 0.003) and FTLD-TDP (F = 6.18, p = 0.0016) but not ALS cases (F = 1.57, p = 0.21). FTLD-TDP misclassified as ALS tended to be of a younger age at death, while LATE-NC cases misclassified as either FTLD-TDP or ALS tended to be of an older age at death.

### Supplementary Tables and Figures

| | | Score Probability | | | |
|---|---|---|---|---|---|
| | | 0 | 1 | 2 | 3 |
| Neuropathological Rating | 0 | 0.88 | 0.12 | $2.95 \times 10^{-4}$ | $1.34 \times 10^{-8}$ |
| | 0.5 | 0.50 | 0.50 | $9.08 \times 10^{-3}$ | $3.04 \times 10^{-6}$ |
| | 1 | 0.11 | 0.79 | 0.11 | $2.64 \times 10^{-4}$ |
| | 2 | $2.64 \times 10^{-4}$ | 0.11 | 0.79 | 0.11 |
| | 3 | $1.34 \times 10^{-8}$ | $2.95 \times 10^{-4}$ | 0.12 | 0.88 |
| | Missing | 0.25 | 0.25 | 0.25 | 0.25 |

**Table S1:** Converting neuropathological ratings to score probabilities

| | ALS | FTLD-TDP | LATE-AD+ | LATE-AD- |
|---|---|---|---|---|
| **Secondary NPDx (%)** | | | | |
| **AD** | 0.40 | 0.39 | N/A | 0.45 |
| **ALS** | N/A | 0.06 | 0.00 | 0.00 |
| **CVD** | 0.03 | 0.02 | 0.03 | 0.01 |
| **FTLD-TDP** | 0.06 | N/A | 0.00 | 0.00 |
| **Hip Scl** | 0.01 | 0.07 | 0.01 | 0.01 |
| **LBD** | 0.04 | 0.10 | 0.59 | 0.00 |
| **PART** | 0.21 | 0.17 | 0.00 | 0.00 |
| **Tauopathy** | 0.01 | 0.07 | 0.02 | 0.04 |
| **Other** | 0.01 | 0.02 | 0.02 | 0.03 |
| **None** | 0.23 | 0.11 | 0.34 | 0.45 |
| **Genetic Mutation (%)** | | | | |
| **C9orf72** | 0.18 | 0.25 | 0.00 | 0.00 |
| **GBA** | 0.00 | 0.00 | 0.01 | 0.04 |
| **GRN** | 0.00 | 0.16 | 0.00 | 0.00 |
| **TBK1** | 0.01 | 0.02 | 0.00 | 0.00 |
| **Trem2** | 0.00 | 0.00 | 0.04 | 0.00 |
| **Other** | 0.01 | 0.03 | 0.00 | 0.01 |
| **None** | 0.80 | 0.53 | 0.95 | 0.94 |
| **Clinical Phenotype (%)** | | | | |
| **AD** | 0.00 | 0.13 | 0.77 | 0.09 |
| **ALS** | 0.91 | 0.02 | 0.00 | 0.01 |
| **bvFTD** | 0.00 | 0.38 | 0.01 | 0.01 |
| **CBS** | 0.00 | 0.05 | 0.03 | 0.03 |
| **DLB/PD/PDD** | 0.01 | 0.00 | 0.04 | 0.61 |
| **FTLD-NOS** | 0.01 | 0.24 | 0.03 | 0.03 |
| **MCI** | 0.00 | 0.00 | 0.01 | 0.00 |
| **MSA** | 0.00 | 0.01 | 0.00 | 0.01 |
| **Normal** | 0.00 | 0.00 | 0.03 | 0.03 |
| **Other MND** | 0.05 | 0.01 | 0.00 | 0.00 |
| **PCA** | 0.00 | 0.00 | 0.00 | 0.00 |
| **PPA** | 0.01 | 0.13 | 0.02 | 0.01 |
| **PSP** | 0.00 | 0.02 | 0.00 | 0.07 |
| **Vascular** | 0.00 | 0.01 | 0.02 | 0.00 |
| **Other** | 0.01 | 0.02 | 0.03 | 0.09 |

**Table S2:** Detailed breakdown of cases included in this study, including incidence of secondary neuropathological diagnosis (NPDx) and incidence of genetic mutations, as well as antemortem clinical phenotype. AD = Alzheimer's disease; ALS = Amytrophic Lateral Sclerosis; bvFTD = behavioral variant frontotemporal dementia; CBS = Corticobasal syndrome; CVD = Cerebrovascular disease; DLB = Dementia with Lewy bodies; FTLD-TDP = frontotempral lobar dementia caused by TDP-43; FTLD-NOS = frontotemporal lobar dementia, subtype not otherwise specified; Hip-Scl = Hippocampal Sclerosis; LATE-NC = Limbic-predominant age-related TDP-43 encephalopathic neuropathological change; LBD = Lewy body disease; MCI = Mild cognitive impairment; MND = Motor neuron disease; MSA = Multiple systems atrophy; Normal = No cognitive impairment; PART = Primary age-related tauopathy; PCA = posterior cortical atrophy; PD = Parkinson's disease; PDD = Parkinson's disease dementia; PPA = primary progressive aphasia; PSP = progressive supranuclear palsy; Tauopathy = Other primary tauopathy; Vascular = Vascular dementia

| ML | ALS | FTD-TDP | LATE-NC | Avg. | Weighted Avg. |
|---|---|---|---|---|---|
| **precision** | 0.862 | 0.885 | 0.852 | 0.866 | 0.862 |
| **recall** | 0.862 | 0.611 | 0.966 | 0.813 | 0.859 |
| **f1-score** | 0.862 | 0.723 | 0.905 | 0.830 | 0.853 |
| **support** | 138 | 126 | 291 | 555 | |
| **accuracy** | | | | 0.859 | 0.813* |

| LR | ALS | FTLD-TDP | LATE-NC | Avg. | Weighted Avg. |
|---|---|---|---|---|---|
| **precision** | 0.873 | 0.868 | 0.970 | 0.904 | 0.923 |
| **recall** | 0.910 | 0.853 | 0.962 | 0.908 | 0.925 |
| **f1-score** | 0.843 | 0.887 | 0.978 | 0.903 | 0.923 |
| **support** | 42 | 38 | 87 | 167 | |
| **accuracy** | | | | 0.923 | 0.903* |

**Table S3**: Cross-validation classification accuracy of maximum-likelihood SuStaIn model (ML) and optimized logistic regression (LR) model including age and SuStaIn stage in predicting pathological diagnosis. The * indicates the metric is balanced accuracy, which is calculated differently from the weighted average (Avg.).

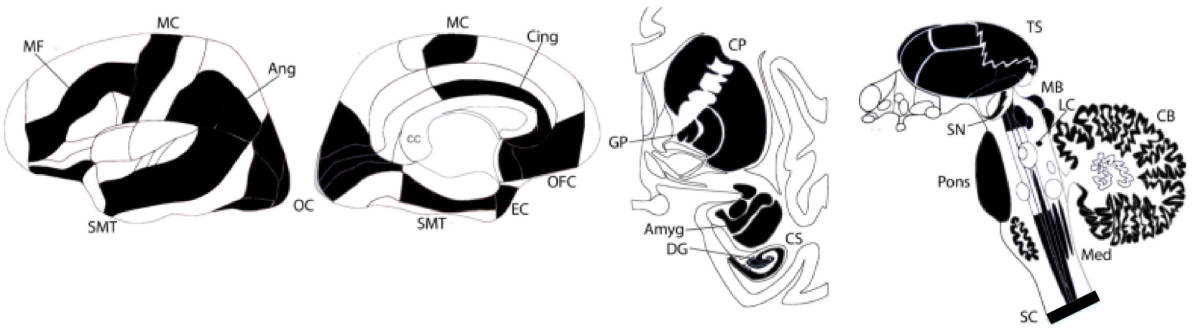


**Figure S1:** Regions sampled for TDP-43 burden in this study are shown in black and indicated with labels. Amyg = Amygdala; Ang = Angular Gyrus;  CB = Cerebellum; Cing = Anterior Cingulate; CP = Caudate/Putamen; CS = CA1/Subiculum; DG = Dentate Gyrus; EC = Entorhinal Cortex; GP = Globus Pallidus; LC = Locus Coeruleus; MB = Midbrain; MC = Motor Cortex; MF = Middle Frontal Gyrus; Med = Medulla; OC = Occipital Cortex; OFC = Orbitofrontal Cortex; SC = Spinal Cord; SMT = Superior and Middle Temporal Gyrus; SN = Substantia Nigra; TS = Thalamus.

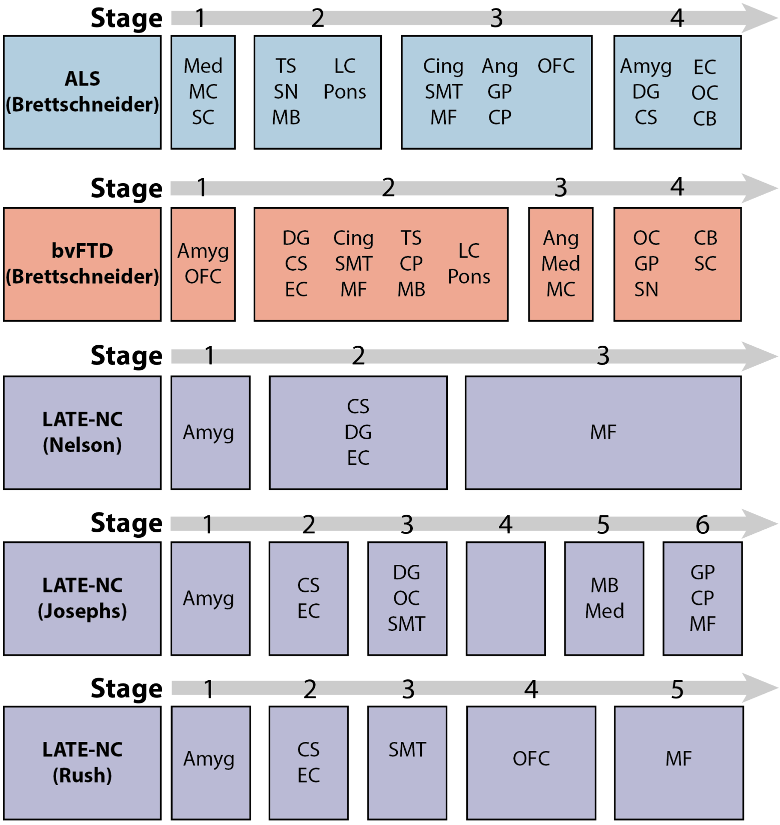


**Figure S2**: Mapping of ROIs included in this study to classical disease stages, as described in Brettschneider et al., 2013 Ann Neurol (ALS), Brettschneider et al., 2014 Ann Neurol (bvFTD), and Nelson et al., 2019 Brain (LATE). Amyg = Amygdala; Ang = Angular Gyrus; CB = Cerebellum; Cing = Anterior Cingulate; CP = Caudate/Putamen; CS = CA1/Subiculum; DG = Dentate Gyrus; EC = Entorhinal Cortex; GP = Globus Pallidus; LC = Locus Coeruleus; MB = Midbrain; MC = Motor Cortex; MF = Middle Frontal Gyrus; Med = Medulla; OC = Occipital Cortex; OFC = Orbitofrontal Cortex; SC = Spinal Cord; SMT = Superior and Middle Temporal Gyrus; SN = Substantia Nigra; TS = Thalamus

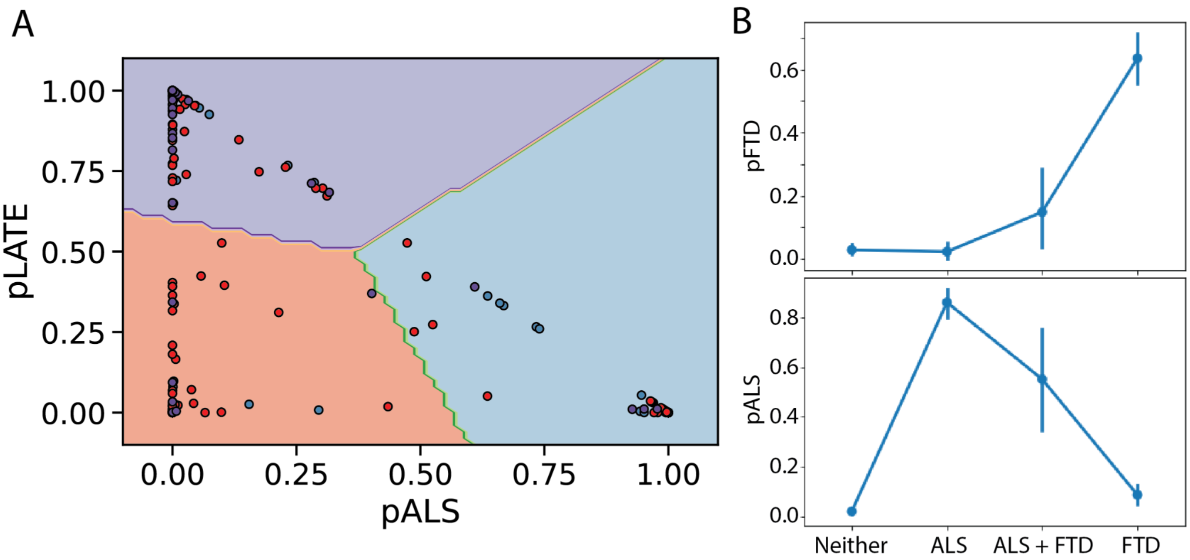


**Figure S3.** Classification decisions. **A**) Visualisation of the decision boundaries for the logistic regression model using SuStaIn-based diagnostic probabilities to classify TDP-43 pathological diagnosis (LATE-NC, FTLD-TDP, ALS). Dot colours represent the true pathological diagnosis of each individual (Blue = ALS, Purple = LATE-NC, Red = FTLD-TDP). Plane colours represent the latent area of classification for each diagnosis. **B**) A subset of subjects was identified that had primary neuropathological diagnosis of ALS with a secondary neurpathological diagnosis of FTLD-TDP, or vice versa (ALS+FLTD-TDP). The SuStaIn-derived classification probability of this group was compared to individuals with a primary diagnoses of ALS or FTLD-TDP without the other as a secondary diagnosis, as well as individuals with neither ALS nor an FTLD-TDP diagnosis. No matter the primary diagnosis, ALS+FTLD-TDP individuals tended to have intermediate probabilities, indicating sensitivity of the SuStaIn algorithm to this apparent mixture of pathological expression patterns.

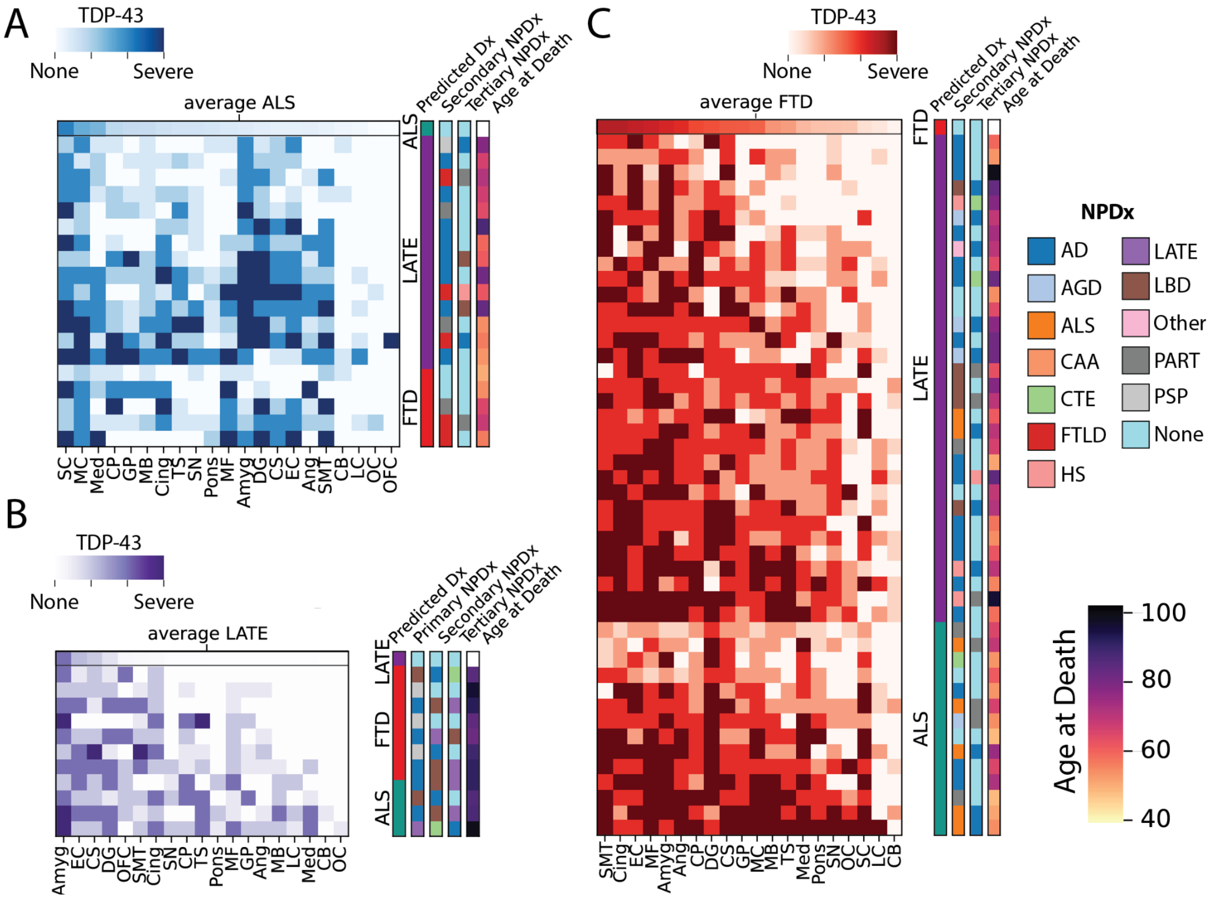


**Figure S4.** Individual regional TDP-43 patterns of misclassified cases. Heatmaps show regional TDP-43 pathology for ALS (A), LATE-NC (B) and FTLD-TDP (C) patients that were misclassified by SuStaIn. The first row of each heatmap represents the average regional TDP-43 pattern across all correctly classified individuals. Heatmaps are otherwise sorted first by predicted (incorrect) diagnosis, and then by overall TDP-43 pathology. Note that a 0 (light color) in the heatmaps denote no pathology OR missing values. Colorbars adjacent to each heatmap show the predicted diagnosis (Dx), secondary and tertiary neuropathological diagnosis (NPDx) and age at death. Since the LATE-NC group is composed of individuals with multiple neuropathologic diagnoses, the primary NPDx is also indicated for this group.

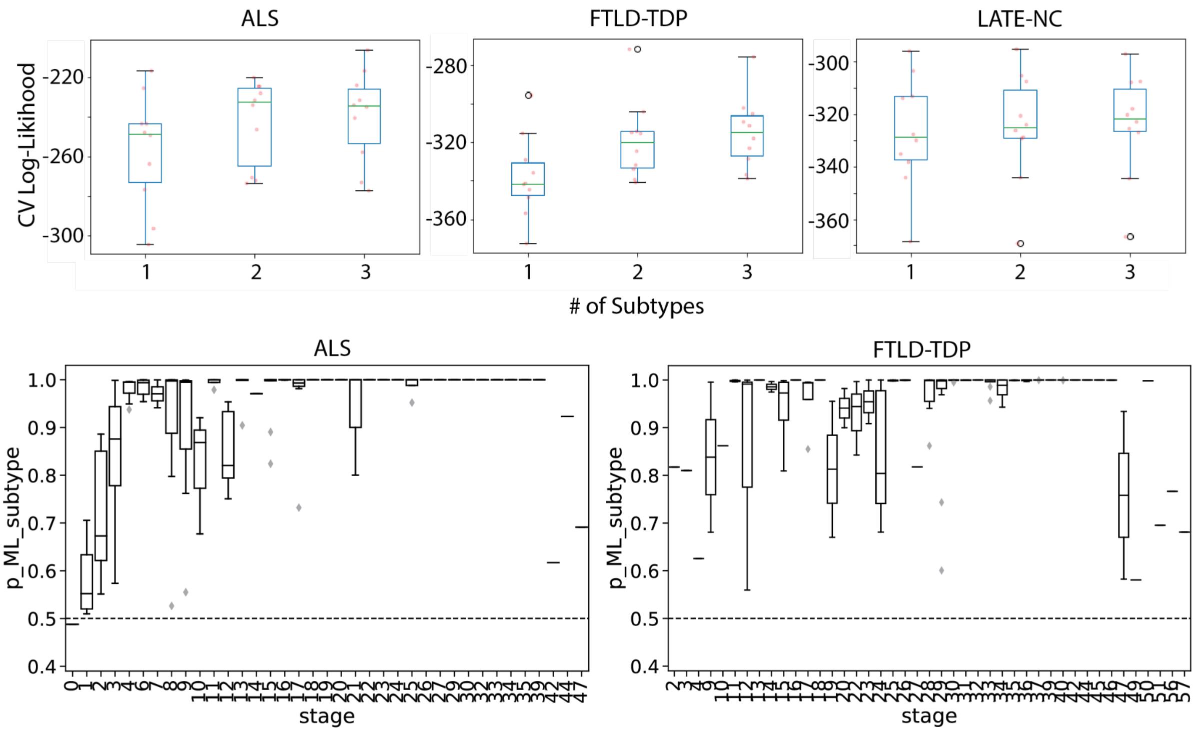


**Figure S5.** Selecting the number of subtypes. (Top) For each TDP-43 diagnostic group, SuStaIn was run using 1,2 and 3 subtype models. To evaluate the number of subtypes, cross- validation was employed and the average log-likelihood was calculated on left-out sample. Each dot represents the mean sample log-likelihood across a given fold. Boxplots were heuristically evaluated, and were used to establish that a 2-subtype model better fits the ALS and FTLD-TDP data, but a 1-subtype model best fits the LATE-NC data. (Bottom) Boxplots representing individual subtype probability across SuStaIn stage is plotted for ALS and FTLD-TDP individuals. Subtype probability was confidently above chance (dotted line) across all non-zero SuStaIn stages, indicating low likelihood of ”cross-over events”.

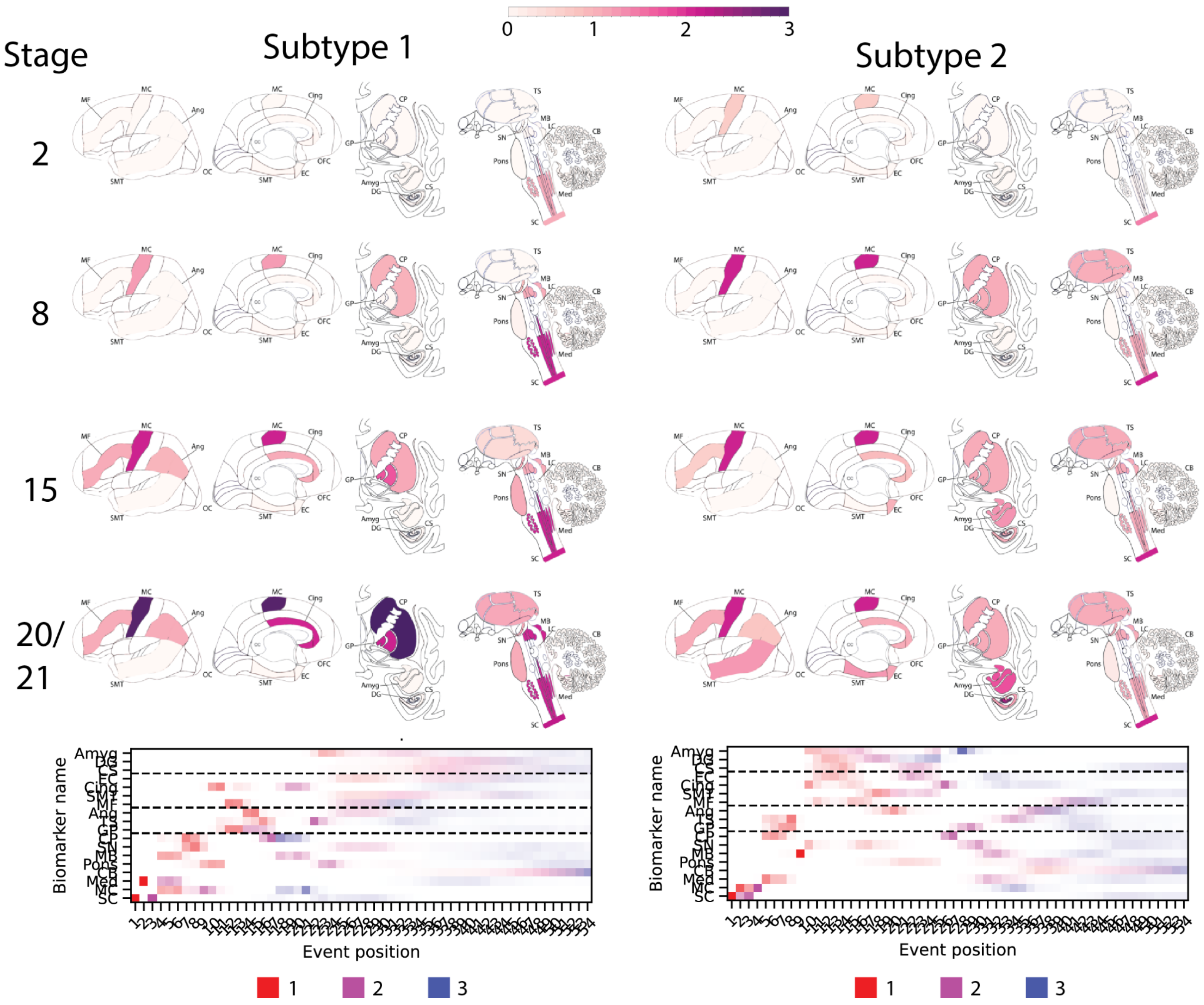


**Figure S6.** Inferred trajectory of regional TDP-43 progression based on individuals with a primary pathological diagnosis of ALS. (Top left) Progression for model combining all ALS patients. Only stages with reduced event ordering uncertainty (e.g. those represented by at least 5 individual donors) are shown. Note this is the same set of maps as in Main Text Figure 1. (Top right) Progression across each ALS subtype identified by SuStaIn. Only stages with reduced event ordering uncertainty (e.g. those represented by at least 3 individual donors) are shown. Positional variance diagrams are included at the bottom for reference. See Fig 2 for explanation for further details. See Main Text Table 2 for subtype Ns and comparisons.

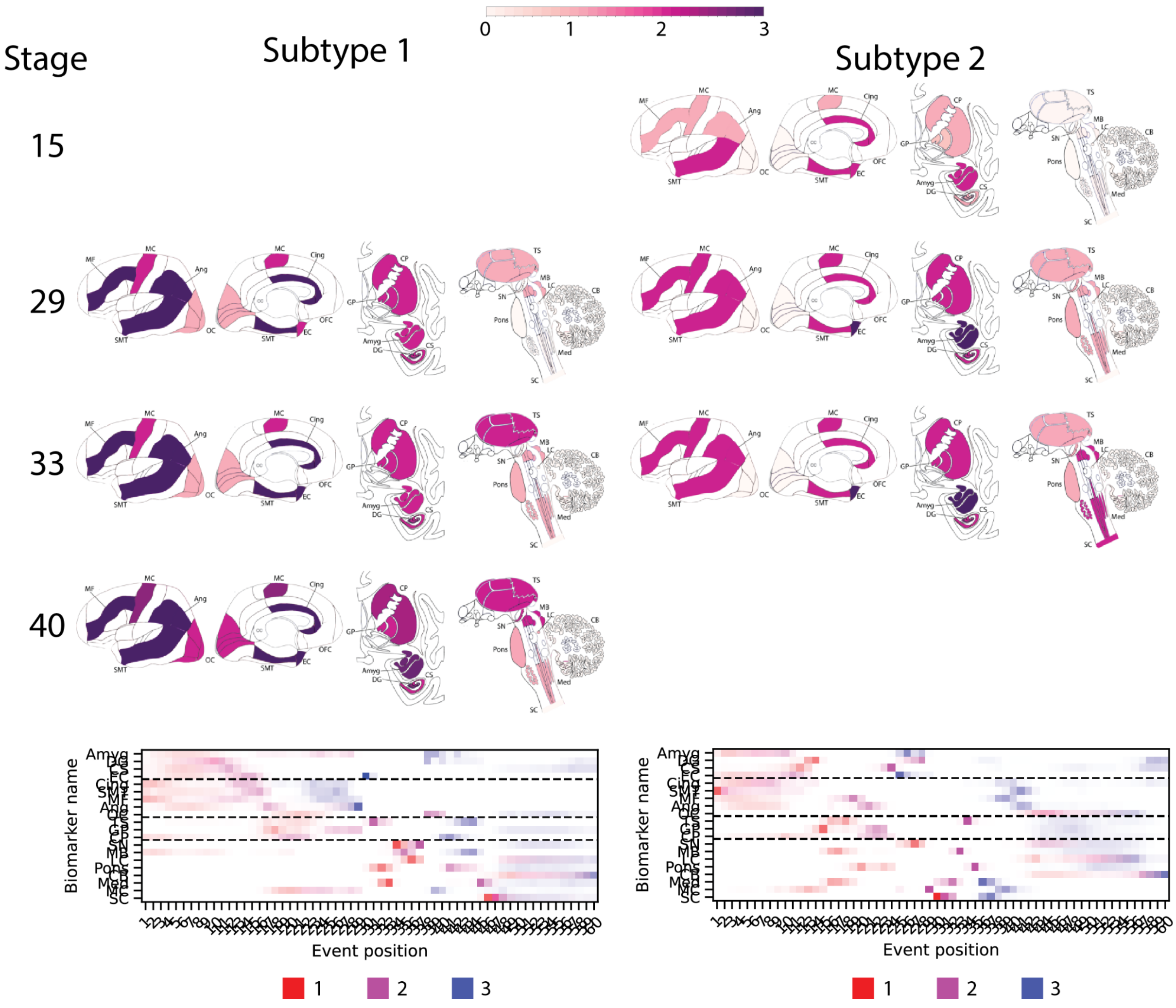


**Figure S7.** Inferred trajectory of regional TDP-43 progression based on individuals with a primary pathological diagnosis of FTLD-TDP. (Top left) Progression for model combining all FTLD-TDP patients. Only stages with reduced event ordering uncertainty (e.g. those represented by at least 5 individual donors) are shown. Note this is the same set of maps as in Main Text Figure 1. (Top right) Progression of across each FTLD-TDP subtype identified by SuStaIn. Only stages with reduced event ordering uncertainty (e.g. those represented by at least 3 individual donors) are shown. The position variance diagram is included at the bottom for reference. See Main Text Table 2 for subtype Ns and comparisons.

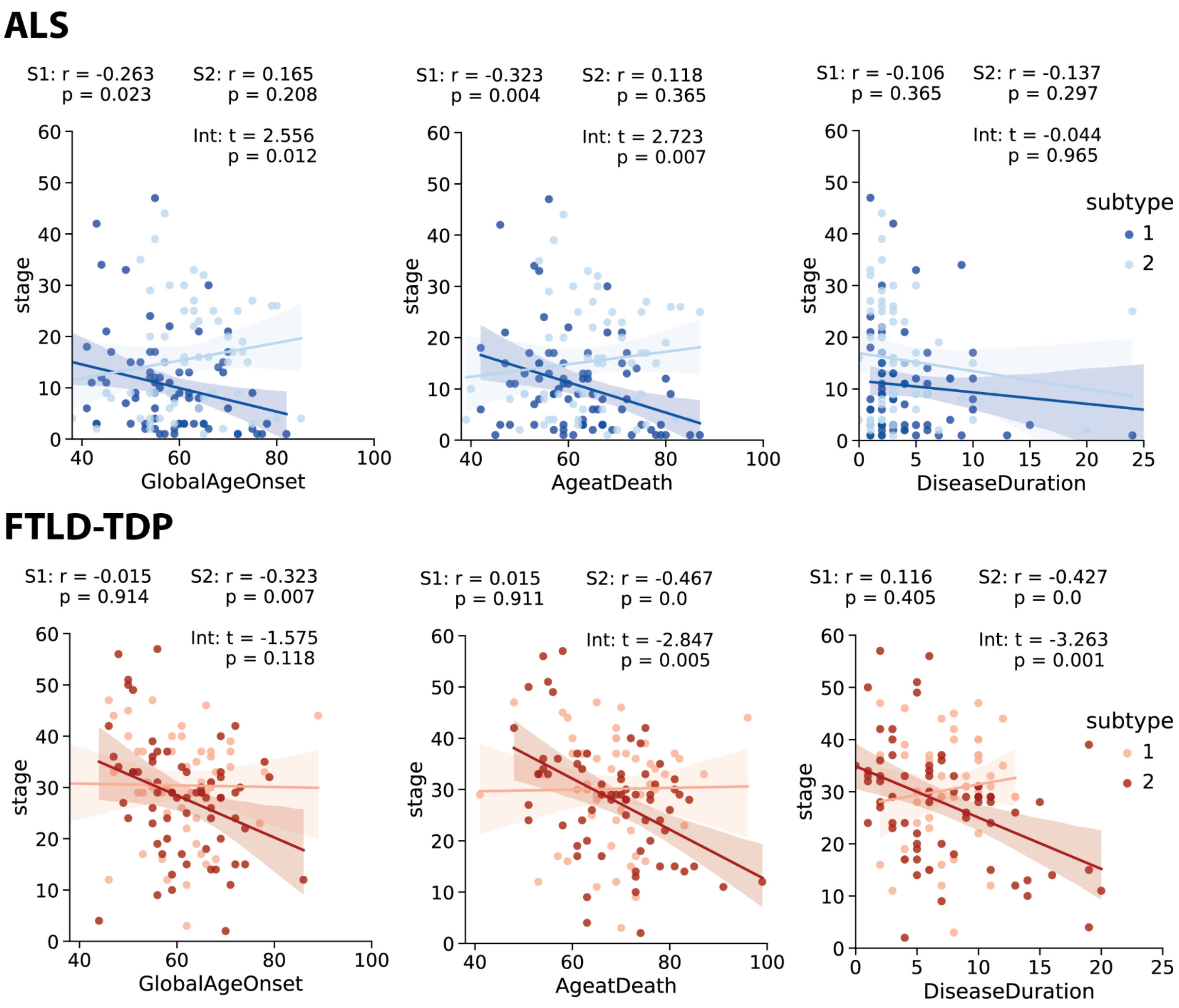


**Figure S8.** The relationship between SuStaIn stage and age at symptom onset (left), age at death (middle) and disease duration (right) are visualized for ALS (top) and FTLD-TDP (bottom) subtypes as in Main Text Fig 2. Statistics demonstrate correlations between these variables separately for each subtype, as well as the interaction between stage and subtype on the variable of interest. Across both pathological entities, the subcortical-predominant subtype (S1 in ALS, S2 in FTLD-TDP) expressed a stronger negative relationship with age and disease duration.
